# Autonomous and policy-induced behavior change during the COVID-19 pandemic: Towards understanding and modeling the interplay of behavioral adaptation

**DOI:** 10.1101/2023.12.09.23299681

**Authors:** Heinrich Zozmann, Lennart Schüler, Xiaoming Fu, Erik Gawel

**Affiliations:** Department Economics, UFZ – Helmholtz Centre for Environmental Research, Leipzig, Germany; Center for Advanced Systems Understanding (CASUS), Görlitz, Germany; Helmholtz-Zentrum Dresden-Rossendorf (HZDR), Dresden, Germany; Research Data Management - RDM, UFZ – Helmholtz Centre for Environmental Research, Leipzig, Germany; Department Monitoring and Exploration Technologies, UFZ – Helmholtz Centre for Environmental Research, Leipzig, Germany; Leipzig University, Institute for Infrastructure and Resources Management, Leipzig, Germany

## Abstract

Changes in human behaviors, such as reductions of physical contacts and the adoption of preventive measures, impact the transmission of infectious diseases considerably. Behavioral adaptations may be the result of individuals aiming to protect themselves or mere responses to public containment measures, or a combination of both. What drives autonomous and policy-induced adaptation, how they are related and change over time is insufficiently understood. Here, we develop a framework for more precise analysis of behavioral adaptation, focusing on confluence, interactions and time variance of autonomous and policy-induced adaptation. We carry out an empirical analysis of Germany during the fall of 2020 and beyond. Subsequently, we discuss how behavioral adaptation processes can be better represented in behavioral-epidemiological models. We find that our framework is useful to understand the interplay of autonomous and policy-induced adaptation as a “moving target”. Our empirical analysis suggests that mobility patterns in Germany changed significantly due to both autonomous and policy-induced adaption, with potentially weaker effects over time due to decreasing risk signals, diminishing risk perceptions and an erosion of trust in the government. We find that while a number of simulation and prediction models have made great efforts to represent behavioral adaptation, the interplay of autonomous and policy-induced adaption needs to be better understood to construct convincing counterfactual scenarios for policy analysis. The insights presented here are of interest to modelers and policy makers aiming to understand and account for behaviors during a pandemic response more accurately.

## 1. Introduction

The COVID-19 pandemic has provided abundant evidence that human behaviors are essential drivers of transmission dynamics, including the course and the duration of outbreaks [1]. Almost all nations implemented public policies aiming to prevent or reduce the spread of the contagion [2]. Such non-pharmaceutical interventions (NPIs), ranging from public information campaigns to stay-at-home-orders, have resulted in significant behavioral changes (here referred to as *policy-induced adaptation*). Numerous research articles and meta studies [e.g., 2, 3, 4, 5] have been dedicated to the question which NPIs are most effective in altering behaviors, with results varying dependent on a range of contextual factors. Particularly relevant have been compliance levels in the population [5]: Even the strictest public mandates (e.g., contact bans) only take effect if a sufficient share of the population chooses to adjust their behaviors accordingly. Beyond “following the rules”, there is convincing evidence [6–8] that individuals change behaviors voluntarily to protect themselves or others against a perceived health threat (here referred to as *autonomous adaptation*). Such behaviors have, for instance, been observed in the early pandemic based on mobility data, when individuals reduced physical contacts and time outside their home prior to this being required by public measures [6].

Behavioral adaptation in a pandemic, its determinants and changes over time need to be understood more precisely. A key question, for example, is in which ways and to which extent public mandates influence individual decisions. It can be quite challenging to establish whether an observed behavior change should be attributed to self-protection or the effect of NPIs, or a combination of both. The relative importance and relationship of autonomous and policy-induced adaptation thus warrants more attention. A number of studies [6–13] has empirically differentiated between voluntary and mandated behavioral response, providing highly valuable insights but also establishing strongly diverging effect sizes (more details in Section 2). However, existing studies focus almost exclusively on the early weeks of the pandemic. It is unclear whether their insights on behavioral adaptation hold over longer periods of time. Given that fear and uncertainty were high in the early pandemic, it is plausible that both autonomous and policy-induced adaptation changed significantly later on, due to emerging issues such as fatigue and non-compliance [14] or a habituation to infection risk in parts of the population [15, 16]. Furthermore, previous work has not accounted for interrelations between autonomous and policy-induced adaptation: The communication and activities of the government, for example, may create awareness or increase public perception of infection risks [10, 17], prompting autonomous risk management. This paints a complex picture, where behaviors evolve dynamically over time, determined by an interplay of autonomous and policy-induced adaptation.

Disentangling this complexity is challenging but there is a strong necessity for it. Acknowledging that human behavior drives disease transmission, we need to be able to better describe and explain behavioral patterns observed in specific situations or over the long term, such as loss of trust in government or diminishing risk perceptions. Furthermore, to identify effective intervention strategies and develop counterfactual scenarios, it is essential to understand which behavior changes result from policies, which changes occur autonomously and whether there are interactions. This could also be useful for behavioral-epidemiological models and thus result in improved decision support for policy-makers. With this article, we contribute to this end in three related steps:

1. We develop a framework for a more precise analysis of autonomous and policy-induced adaptation by synthesizing various literature on human behavior during the COVID-19 pandemic. We briefly characterize both adaptation mechanisms, their determinants, their relationship and potential changes over the course of the pandemic.
2. We carry out an empirical investigation of both role and relevance of autonomous and policy-induced adaptation in Germany using a variety of publicly available data. The analysis addresses the “second wave” of the pandemic (fall of 2020) and longer-term trends affecting behavioral adaptation (diminishing risk perceptions & eroding compliance).
3. We give an overview of how autonomous and policy-induced adaptation have been represented in behavioral-epidemiological models and discuss how empirical and conceptual models may be further improved.

The paper is organized as follows: In the next Section, we give an overview of related literature and relevant gaps. In Section 3, we develop and present our framework, followed by an empirical analysis of the German case in Section 4. Subsequently, we discuss the current state and promising directions for model-based analysis of behavioral adaptation (Section 5). We then discuss the insights and limitations of our analysis (Section 6). Section 7 concludes.

## 2. Background: Autonomous and policy-induced adaptation during the COVID-19 pandemic

The COVID-19 pandemic has changed human behaviors in various ways, (e. g. travel/mobility and social practices, consumption patterns). Here, we focus on behaviors relevant for the *transmission* of the virus, including physical contacts, mobility patterns, the use of preventative measures such as testing and facial covering, and more (cf. *Note 1*). Based on the premise that human behaviors are key drivers of transmission [1], we analyze two key forms in which individuals have changed their behaviors during the COVID-19 pandemic: Autonomous and policy-induced adaptation. Synthesizing insights from various bodies of behavioral literature, we first characterize these two adaptation mechanisms and their determinants (Sections 2.1 and 2.2). We then give an overview of existing studies that have disentangled autonomous and policy-induced adaptation empirically (Section 2.3) and outline which aspects of their relationship remain insufficiently understood and will be addressed by our framework.

### 2.1 Autonomous adaptation

Autonomous adaptation refers to the idea that individuals assess the risk that COVID-19 poses to their health, their household or community and adjust their behaviors voluntarily to mitigate this risk, while considering the costs of adaptations. This idea is well-rooted in a number of theories and explanatory frameworks of the social sciences. Psycho-social theories of health behavior [e.g., 18, 19, 20] explain self-protective behavior through perceptions about infection risks, the efficacy of preventative behaviors, and potential barriers to engaging in such behaviors. In economic analyses [e.g., 21, 22, 23], this is often approached as a cost-benefit calculation, assuming that individuals trade-off the risk of an infection against the cost of changing behaviors, which may include foregone income due to social distancing, but also the loss of utility resulting from having fewer social contacts [24].

Evidence for such behavior has been found throughout different phases of the pandemic, for instance when individuals reduce their mobility before restrictions are in place [6, 7, 10, 13, 25] or maintain fewer contacts and stay out of public areas even after these are lifted [26, 27]. Empirically, self-protective behavior has been associated with a number of socio-demographic and attitudinal variables [28, 29]. Perceptions of risks, for example about the severity of an infection [30–33] or of the effectiveness of behavioral adaptations [32, 34] have been found to impact both the number of private contacts as well as compliance with public measures (more on the latter below). [35] have reviewed the literature on risk perceptions and identified recurring predictors for the extent to which COVID-19 is perceived as a health threat. Key demographic factors are age, gender and income and education levels. Relevant personal factors include the physical and mental health state of an individual, their media exposure, knowledge about COVID-19, trust in the government, media and science [35]. In the United States context, different works have also found political preferences relevant for the level of perceived risk [36, 37]. Besides perceived risk or inclination to self-protect, multiple studies provide evidence that circumstances frequently seem to dictate whether it is possible to act on this, for instance when the housing or work situations do not allow effective social distancing [29, 34, 38].

### 2.2 Policy-induced adaptation

Policy-induced adaptation occurs when individuals or groups alter their behavior in response to a specific policy or intervention. Policy-makers have responded to the COVID-19 pandemic with wide range of non-pharmaceutical interventions, spanning local to supra-national levels as well as short-term to long-term periods, with measures as diverse as stay-at-home-orders and the free provision of sanitary equipment [39]. Interventions relevant for transmission prevention (cf *Note 2*) include measures aiming to reduce the number of physical contacts by limiting access to public areas (e.g., school closures) or issuing stay-at-home orders. They also include interventions aiming to reduce the probability of transmission from physical contacts, e.g., through mask-wearing, preventive testing and vaccination. The widespread and heterogeneous use of NPIs during the pandemic has led to a large and growing body of literature dedicated to identifying the most effective and efficient interventions [40–43]. However, there is an increasing awareness that there are no one-size-fits-all solutions with respect to NPIs, as a number of framework conditions determine their successful application [5, 44, 45].

As most NPIs require a sufficiently large share of the population to adhere to them, their success is determined by the behavioral response or degree of *compliance* in the population [29]. Even strict behavioral mandates require individuals willing to comply and are premised on controls, sanctions and an understanding of their necessity, which is frequently not considered when the effectiveness of NPIs is evaluated. Nonetheless, the determinants of compliance have been examined by a number of studies across nations and within populations [e.g., 46, 47, 48]. It is difficult to generalize findings, as these strongly depend on the type of NPI, as well as situational, economic and cultural factors [47, 49]. Surveys of self-reported compliance consistently suggest that female respondents are more likely to report compliant behavior [e.g., 48, 50] whereas the impact found for age, income and education levels varies. Perceived social norms have been found to strongly impact compliance levels [46, 51] as well as perceptions about the risk of an infection, or the efficacy of government response measures [48, 52, 53]. Non-compliance, on the other hand, was found among those exhibiting lower trust in government, lower empathy, science skepticism and conspiratorial beliefs [54–57]. From an economic perspective, the degree of compliance to behavioral mandates is generally perceived as subject to individual choice weighing the costs and benefits that arise [58].

Compliance is furthermore dependent on the implementation strategy, which may make use of diverse instruments, ranging from coercion to persuasion to incentivization [59]. Interventions based on coercion tend to be harder to circumvent, if they are enforceable [60]. However, they are associated with significant economic and political cost due to infringements on individual liberty [61, 62]. Persuasion or incentivization, on the other hand, leave more room for non-compliant behavior to occur and are thus predicated on trust [14, 63–65]. They are less likely to be perceived as intrusive and thus may spark less aversion [66], for instance when income support allows individuals to follow stay-at-home orders by mitigating their financial losses [67].

### 2.3 Disentangling autonomous and policy-induced adaptation

Disentangling autonomous and policy-induced adaptation is challenging because the motivations for an individual’s behavior, their perceptions and attitudes are difficult to infer from available data. In the early weeks of the pandemic, a number of studies have differentiated between “voluntary” and policy-induced behavioral adaptation [6–13]. Analyzing changes in mobility patterns before and after the implementation of lockdowns, these studies find significant effects of both autonomous response and policy mandates, albeit with diverging effect sizes. Examining data on visits to commercial establishments, [12], for instance, found that “much of the decline in foot traffic early in the pandemic was due to private precautionary behavior”(p. 1). [8] find both effects to be in the “same order of magnitude” (p. 874), with autonomous and policy-induced adaptation reducing deaths by 9% and 14%, respectively. Other studies acknowledge that substantial reductions in contacts occurred due to autonomous adaptation but see these changes as “significantly smaller without a lockdown in place” [11] (p.2) or “not sufficient to bring the R number below one” [9] (p.30).

These studies made valuable contributions to the understanding of autonomous and policy-induced adaptation. To empirically disentangle both, however, they assumed that these are separate effects. Most studies compared pre- and post-lockdown behaviors, while other NPIs such as public information campaigns and behavioral recommendations were already implemented. Thus “voluntary” behavior change rather refers to the absence of mandates to shelter in place than to an absence of policies in general [8]. As we will address in more detail below, it is likely that both adaptation mechanisms are in a more complex relationship, for example when activities of the government and public debate about NPIs enhance the perception of risk among individuals. Moreover, almost all existing studies of autonomous and policy-induced adaptation stem from the very beginning of the pandemic. Given that their determinants (see Sections 2.1 & 2.2) changed over time, their relationship likely varies over the course of a dynamically unfolding pandemic. Hence, a more precise framework is needed to understand the interplay between autonomous and policy-induced adaptation.

## 3. A framework for analyzing behavioral adaptation: Confluence, interactions and time variance

In this section, we present an analytical framework focused on the interplay of autonomous and policy-induced adaptation. Fig 1 illustrates key components of this framework: Autonomous and policy-induced adaptation are at the center of this perspective, which assumes that individuals change their behavior based on an analysis of cost and benefit that considers self-protection and non-pharmaceutical interventions. Drawing from the literature presented in the previous section, we assume that this process is influenced by a range of demographic and personal factors, social norms, cultural beliefs, and the information available to the individual. While determinants such as socio-demographic background or social norms have received considerable attention elsewhere [e.g., 35, 46, 50, 51], we focus here on the relationship between these two adaptation mechanisms. Considering behavioral adaptation as a “moving target”, we explore three key phenomena of the interplay between autonomous and policy-induced adaptation, indicated in color in Fig 1:

1. *Confluence*: Autonomous and policy-induced adaptation can overlap, for example when a high propensity for self-protection results in behavior that is compliant with existing mandates. However, they may also diverge, which can result in a number of distinct effects for overall adaptation (see Section 3.1). In Fig 1, this is illustrated as two blue circles, which overlap to a varying extent.
2. *Interactions:* Autonomous and policy-induced adaptation are subject to a variety of interactions, for example when non-pharmaceutical interventions increase risk awareness and thus prompt higher self-protection. In Section 3.2, we address such interactions with a focus on risk signals as well as the role of trust in their processing and compliance with NPIs. In Fig 1, the interactions considered here are marked in red.
3. *Time variance:* Due to variations in their determinants, the interplay between autonomous and policy-induced adaptation changes over time. In Section 3.3, we substantiate this by addressing changes in two crucial variables over time (risk perception and trust). Fig 1 illustrates time variance through the green arrows at the bottom.

**Fig 1.**
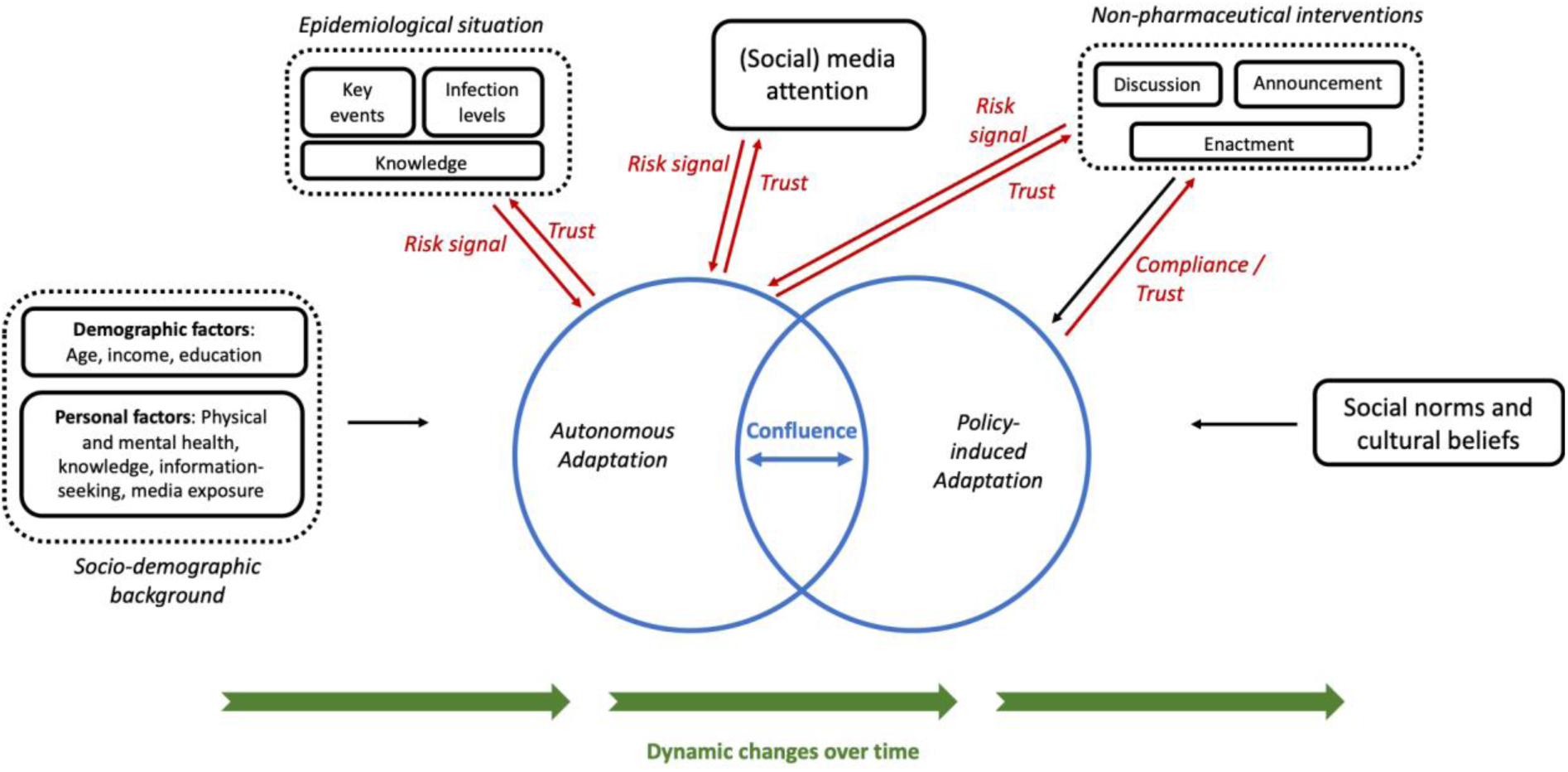
Conceptual framework of behavioral adaptation. The framework focuses on three key phenomena of the interplay of autonomous and policy-induced adaptation: Confluence (blue), interactions (red) and time variance (green).

### 3.1 Confluence

Confluence, here, refers to the process in which autonomous and policy-induced adaptation mechanisms combine and form the actual and observable behavior of an individual. Our framework assumes that this process includes overlaps as well as divergences. Intuitively, it would seem straightforward that both adaptation mechanisms are complementary, i.e., that a high propensity to self-protect predicts a high degree of compliance with NPIs [53, 68]. This is substantiated by the significant overlap in their determinants (cf. Sections 2.1 & 2.2), including demographic factors and the perceived risks of infection. However, the will to self-protect is not always aligned with the objectives of containment policies, particularly in a heterogenous population with diverging (perceived and actual) risks of infection and costs of behavioral change. When both come together, a range of results can emerge.

To illustrate this, we present four simplified cases in Fig 2, juxtaposing an individual propensity for behavioral adaptation (to mitigate risk for one’s health) with a “mandated adaptation”, i.e., behavior change required by NPIs. First, consider the two cases on the left side, where the behavioral adaptation conforms to the policy objectives. In the top left case, this is the result of a low(er) propensity to self-protect combined with “compliant” behavior, which may be due to altruistic or prosocial motivations [69, 70], the fear of penalty [71] or social deviance. In the bottom left case, a strong individual propensity for adaptation exceeds what is required by NPIs, resulting in a form of “overcompliance” or use of preventive behaviors beyond the mandated [e.g., 25, 26]. Second, we consider cases where behavioral adaptation falls short of policy objectives. Here, as well, different plausible explanations exist. In the top right example, non-compliance results from a low(er) individual propensity for adaptation combined with an objection to the current set of rules. And even in cases where individual propensity to adapt is high (bottom right), circumstances may prevent individuals from complying with NPIs, for example due to their occupation, housing or sanitary conditions [17, 65, 72, 73].

**Fig 2:**
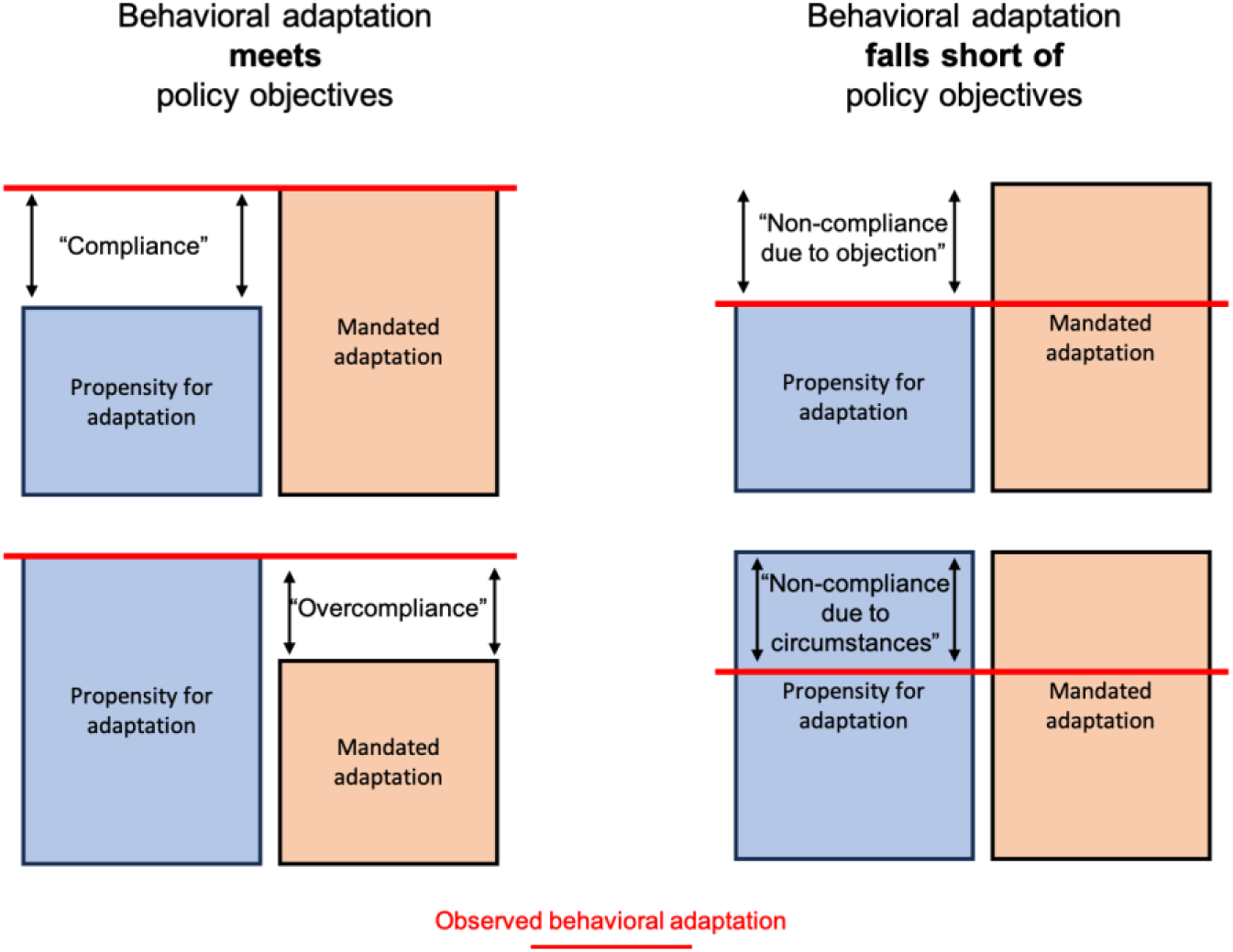
Alternative explanations for observed behavioral adaptations. In each of the four examples, the red line represents an empirically observable behavior (e.g., number of physical contacts), while the boxes indicate individual propensity for behavioral adaptation (blue) and an assumed mandated level of adaptation (orange). Note that behaviors may fall within a spectrum between the depicted cases, for instance if observed behavioral adaptation exceeds individual propensity to adapt but falls short of mandates. Also note that there may be cases of inadvertent non-compliance or overcompliance, for instance when individuals are unsure about the current set of “rules”.

Hence, autonomous and policy-induced adaptation may overlap or diverge, but which concrete, observable behavior results from their confluence is not obvious. This presents a challenge, as commonly used data (e.g., on mobility) are inadequate to understand such subtleties and detailed time series on attitudes and motivations are frequently unavailable. The question whether observed behavioral changes are the result of autonomous or policy-induced adaptation, or a combination of both, has therefore not been studied sufficiently. Nonetheless, it is highly relevant to understand these patterns more precisely: As [13] point out, if policy action crowds out voluntary efforts, then “mandates achieve the outcome at a greater cost” (p. 2). And, as we address in more detail below, strict mandates can have negative impacts on social cohesion and trust over time [66]. Thus, better understanding the confluence of autonomous and policy-induced adaptation is relevant for analyzing and modelling behavioral patterns in a pandemic, which will be discussed further in Section 5.

### 3.2 Interactions: Risk signals, trust and compliance

While autonomous adaptation in our framework refers to an individual trading off infection risk and the cost of changing behaviors, we do not assume that individuals form their perceptions in isolation. Instead, individuals are assumed to evaluate information available to them, which includes the processing of risk signals they perceive from various sides. Here, we focus on three prominent sources of such risk signals:

- *Government activities*: The government is an important conveyor of information to many, and thus its communication and activities may receive substantial weight in that process. In the early pandemic in particular, there is substantial information asymmetry, as governments tend to have access to data and experts unavailable to the public. Thus, government communication may be an influential signal to individuals’ assessing infection risk: In the United Kingdom, for instance, 71% of respondents in a study reported to have changed their behavior based on government guidance in mid-March 2020 [17]. Studies examining the impact of specific events such as the declaration of emergency at the state level [6, 13] have demonstrated that such signals in particular conveyed “information about the seriousness of the epidemic“ [10] and led to a significant public response. Similarly, the actions taken by the government can be interpreted as a signal, where the stringency of deployed NPIs indicates the severity of the situation. And even before NPIs enter into legal effect, debates and decisions about them can have (unintended) consequences, such as people frequenting public spaces more often before an announced lockdown enters into effect [13]. Besides the timing, the messaging and style of signals has been found to be highly impactful [74].
- *Epidemiological situation*: The current infection levels and the extent to which individuals are aware of them signal a certain level of health risk. It has been shown that this effect is stronger with confirmed cases within one’s own social network [75]. In addition, key events with high visibility can have significant impacts on perceived risk and lead to spikes in media interest [15]. Of key relevance is also knowledge, which we may define loosely as the information, insights, and understanding that an individual possesses about COVID-19 and the extent to which this informs and guides their decisions and behaviors (cf. *Note 3*). Such knowledge may include information about common symptoms, transmission routes, susceptibility, mortality risk and more [76].
- *Media*: Public visibility of the pandemic and its media coverage [77] may also emit highly relevant risk signals. Extensive evidence suggests risk perception and compliance with NPIs can be influenced by exposure to media, particularly social media, and how the COVID-19 pandemic is portrayed in the news [78–80]. Strongly linked to the above-mentioned signals, media coverage impacts the overall frequency and prominence of COVID-19, likely resulting in effects to which individuals consider it relevant (*availability bias*). The perceived severity of the situation may also be impacted by framing of signals, for instance when a greater focus is placed on fear- and anxiety-inducing messages, such as the death toll, rather than highlighting recoveries or positive developments, as subjective emotions often play a stronger role than factual information [65].

Trust plays an important role in the framework presented here. For one, it influences the extent to which risk signals are heeded. If information about the progression and severity of the pandemic is not considered credible, it may not have the desired impact on behaviors. Trust in (social) media reporting, for example, has been linked to the adoption of preventive measures as well as impacts on the overall evaluation of the pandemic [81, 82]. Beyond its role as a mediator for the processing of risk signals, trust in the government and its scientific institutions has been associated with a range of factors including the willingness to change behaviors and adhere to mandates [55, 83, 84] (cf. *Note 4*). The perceived competence and consistency of the government response, as well as the extent to which costs of adaptation are mitigated, affects compliance levels in the population, for example the willingness to self-isolate in case of an infection [65, 67, 85]. Governments enjoying high levels of trust may thus be able to use moral arguments and suasion (“Do the right thing”) with a higher chance of success [65].

### 3.3 Time variance

There is considerable evidence that certain determinants of behavioral adaptation, such as perceived risk of infection, are time-variant. This suggests, in turn, that the dependent variables autonomous and policy-induced adaptation may change significantly over time. Due to their critical role in our framework, we focus here on two key variables: The level of perceived risk as a determinant of both autonomous and policy-induced adaptation and trust as a key mediator of their interactions:

- *Diminishing risk perception*: Empirical evidence suggests that the average perceived risk of infection has declined over time, at least in parts of the population [86]. Particularly in the early weeks of the COVID-19 pandemic, fear and uncertainty tended to be particularly high. As time progresses, the risk perceived from the same epidemiological situation, such as reported new cases, may decline. This is due to a variety of reasons, such as increasing knowledge about the virus and its transmission channels or the increasing ability to manage or reduce the risks of infection or a severe course, e.g., through facial masks or vaccinations. In addition, the constant exposition to the threat of an infection may lose salience for perceived risk over time caused by saturation with the topic, or declined information-seeking as habituation effects set in [15, 50]. [79] have investigated the impact of media coverage on public attention over time and found that despite constantly high news coverage, attention levels for the pandemic were decreasing. A lower risk perception is, in turn, associated with decreased autonomous adaptation and compliance with policies. This is related to the widely discussed notion of “pandemic fatigue”, which is contested as a scientific concept [87]. Nonetheless, large-scale studies have found for behaviors such as physical distancing a decline in adherence levels, suggesting an eroding compliance with at least some aspects of policy over time [14]. Individual longitudinal studies find self-reported compliance to remain high for most participating individuals, where “approximately 15% of participants had decreasing levels of compliance across the pandemic, reporting noticeably lower levels of compliance in the second wave” [50] (p.781). Interestingly, particular behaviors such as staying at home with symptoms have been less adhered to, which has potentially strong impacts on transmission. Some have related decreases in compliance over time also to the idea of “alert fatigue”, i.e., a lack of capacity to understand rules that are frequently changing depending on place and time [88], which may result in cognitive overload and inadvertent non-compliance [89].
- *Erosion of trust*: When considering changes in policy-induced adaptation over time, trust as a moderator of effect strength has high relevance. Evidence suggests that the decline in compliance was lower in countries with initially high levels of trust in government [84]. A variety of studies studying trust over longer periods of the pandemic indicate that trust in government and science agencies have substantially declined [83, 90–92]. There are manifold explanations for this, including political partisanship [92], misinformation [93] and perceived competence and fairness of the government handling of the pandemic [84]. The loss of trust has also been related to the use of certain NPIs which may create “control aversion” [66].

## 4. Empirical application: Behavioral adaptation in Germany during later stages of the pandemic

In this Section, we use our framework to carry out an empirical analysis of autonomous and policy-induced adaptation in Germany. We first empirically investigate behavioral adaptation during the “second wave” of the pandemic (autumn/winter 2020/21), using a similar approach as existing research from the early weeks of the pandemic, thus complementing their works. We then conduct a comparative analysis of interactions between autonomous and policy-induced adaptation, focusing on risk signals and public attention during spring and fall of 2020. Finally, we address time variance of behavioral adaptation by investigating whether (i) risk perceptions diminished and (ii) we find evidence for an erosion of trust and compliance in Germany. For these analyses, we combine a variety of publicly available data, analyze these statistically and contextualize them with qualitative information. Detailed descriptions of methods and data are presented in Supplementary Information S1 and S2.

### 4.1 Interplay of autonomous and policy-induced adaptation in Germany: Spring and fall 2020

Germany’s management of the early pandemic (March-May 2020) is widely regarded as successful, characterized by comparatively low case numbers and death toll. Besides a swift policy response at an early stage, this may be attributed to autonomous adaptation: [8] have analyzed mobility changes for the “first wave” in Germany and twelve other countries, finding both mechanisms of behavioral adaptation to have similar effect sizes. After the first wave subsided, restrictions were lifted gradually and the summer in 2020 was characterized by comparatively few COVID-19 cases.

As our literature overview in Section 2.3 indicates, this assessment of behavioral adaptation aligns with similar studies conducted in various contexts during spring of 2020 [e.g., 7, 13]. However, a less thoroughly studied question is how the interplay of autonomous and policy-induced adaptation evolved during later stages of the pandemic. To address this gap, we examine the situation in the fall of 2020, which paints a different picture. By early October, the number of infections in Germany began to increase rapidly. At this point, NPIs were mostly implemented at the county level with varying degrees of stringency. In early November 2020, a ‘lockdown light’ was enacted on the national level which restricted public events and private meetings but allowed retail shops to remain open. After this had failed to reduce infection levels sufficiently, a full lockdown followed on December 16. We focus our first statistical analysis and the analysis of interactions on this ‘second wave’ which began in October 2020, according to the German center for disease control [94]. We consider a time period until the end of January, 2021, by which the wave had largely subsided and the vaccine roll-out had begun, which likely introduced further changes in behaviors and thus marks a good finishing point [95]. The heterogeneity of response measures and infection levels throughout Germany, the national lockdowns as well as the public perception of COVID-19 make this an interesting comparison to the widely studied first wave.

#### 4.1.1 Data

To empirically investigate the relationship of autonomous and policy-induced adaptation, we follow an approach similar to existing literature [e.g., 8, 13], combining publicly available data from various sources, for which the smallest shared geographical unit available are the 16 German federal states. Key data enabling our analysis are:

- *<u>Changes in mobility</u>*: As the key indicator for human behavior driving transmissions, we consider the average changes in mobility compared to the respective month in 2019 on a given day in each state, which were estimated based on aggregated GPS data stemming from mobile phone devices by the Federal Statistical Office of Germany [96].
- *<u>NPI stringency</u>*: To assess the impact of NPIs, we use a composite policy stringency index, which is conceptually and methodologically similar to the well-known *Oxford COVID-19 Response tracker* [39]. The data for this index was compiled at high resolution by an interdisciplinary team for the *Corona Data Hub Germany* [97] and captures the varying intensity of NPIs deployed in individual states as well as on the national level. Thus, we assume it provides a useful variable to account for policy-induced adaptation.
- *<u>7-day-incidence</u>*: We use the data on the number of infections per 100,000 inhabitants in the past seven days (subsequently referred to as “incidence”) provided by [98], for the state and national level in Germany. We use incidence as a proxy for the risk signals emanating from current infection levels. As incidence numbers were reported daily by national and local media, they are also related to public visibility. In absence of high frequency data directly capturing autonomous adaptation, we consider incidence the best available proxy.

Combining data in daily frequency over four months and 16 federal states results in all overall sample size of n =1968 observations. In Fig 3, the three key variables used in our analysis are plotted for the individual federal states.

**Fig 3.**
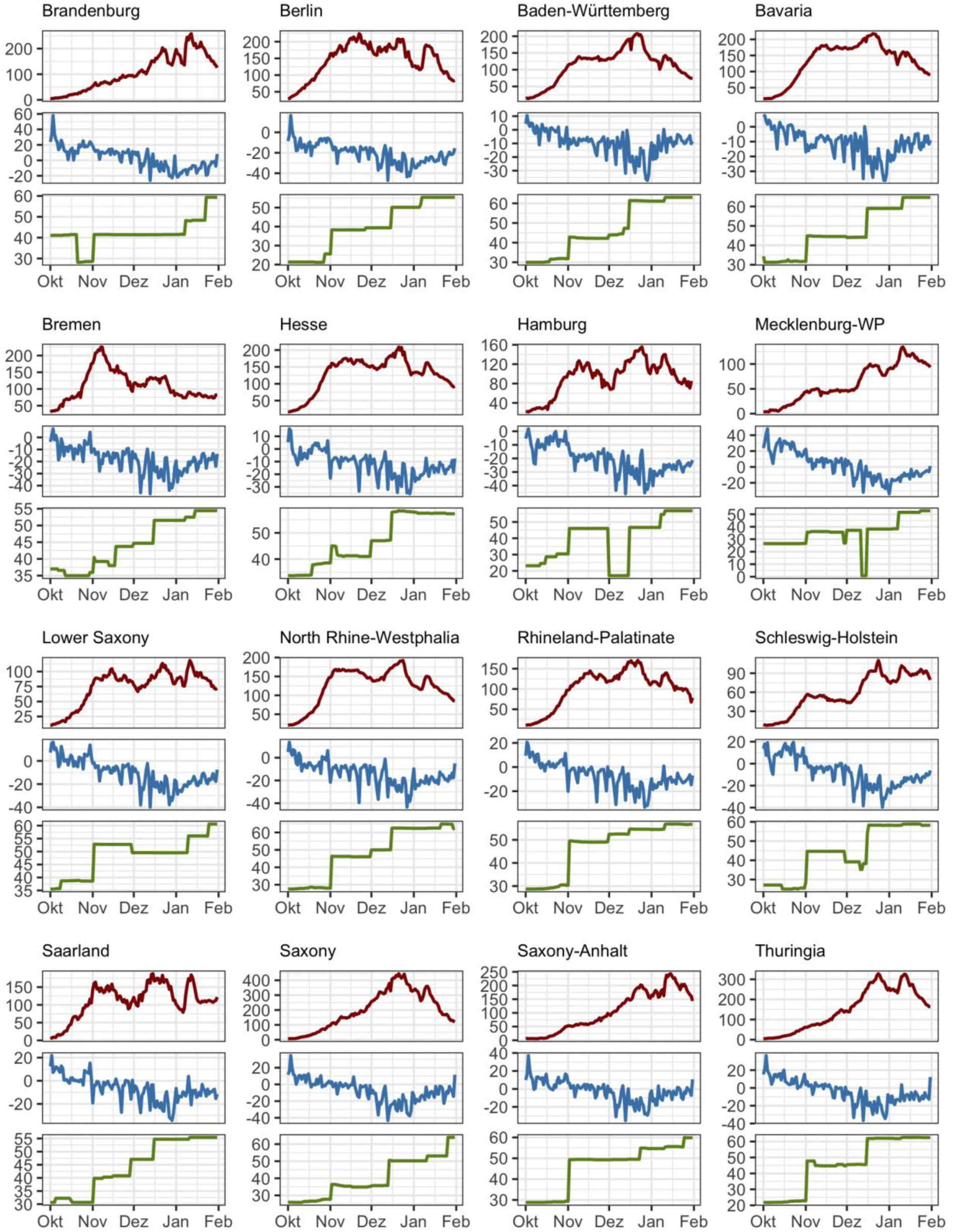
Incidence, mobility changes and policy stringency in Germany. The plot depicts data stemming from [96–98] for the 16 federal states of Germany between Oct 1, 2020 and Jan 31, 2021.

#### 4.1.2 Statistical Analysis

We estimate a range of linear models using fixed effects to account for heterogeneity among the German states. Across all models, the daily average change in mobility is the response variable. Incidence levels as a measure for infection risk are used as predictor, employing the natural logarithm due to at times exponential growth in case numbers. The stringency of policy response is also a predictor across all models, either measured by the state-level stringency index or as an ordinal variable, differentiating three distinct phases of national policy response (local measures, lockdown light, hard lockdown – in dependence of the date). We control for different weekdays as well as daily temperature and precipitation due to the change of season occurring in the studied time span. In Supplementary Information S1, we present detailed model specifications and diagnostics.

Across all models, we find a consistent and significant negative effect of incidence and stringency on mobility (for detailed regression coefficients see S1 Table 1). With a relatively parsimonious approach, the models can capture up to 70% of the variance in the data, with remaining variations likely due to geographical aggregation and, perhaps, seasonal holidays. The results indicate that both autonomous risk management and containment policies resulted in relevant decreases in mobility: Between October 1 and December 24, the average increase in incidence (from 13 to 193 new cases per 100,000 inhabitants) leads to a little more than 20% reduction in mobility, assuming a weekday and holding stringency at its mean value and weather data at the average for the month. The model predicts a reduction of about 9% due to (additional) stringency of policies during the same time span, under the same assumptions and the mean incidence level of the considered period. Interestingly, if policy stringency is included as an ordinal variable describing three phases of national response, slightly more variation in the data can be captured than by using the state-level stringency index, with the impacts of incidence remaining robust. This may be an indication for the significance of national events like the initiation of lockdowns. In Fig 4, we visualize the marginal effects of incidence on mobility under county-level (“local”) measures, as well as the national “lockdown light” and a hard lockdown.

**Fig 4.**
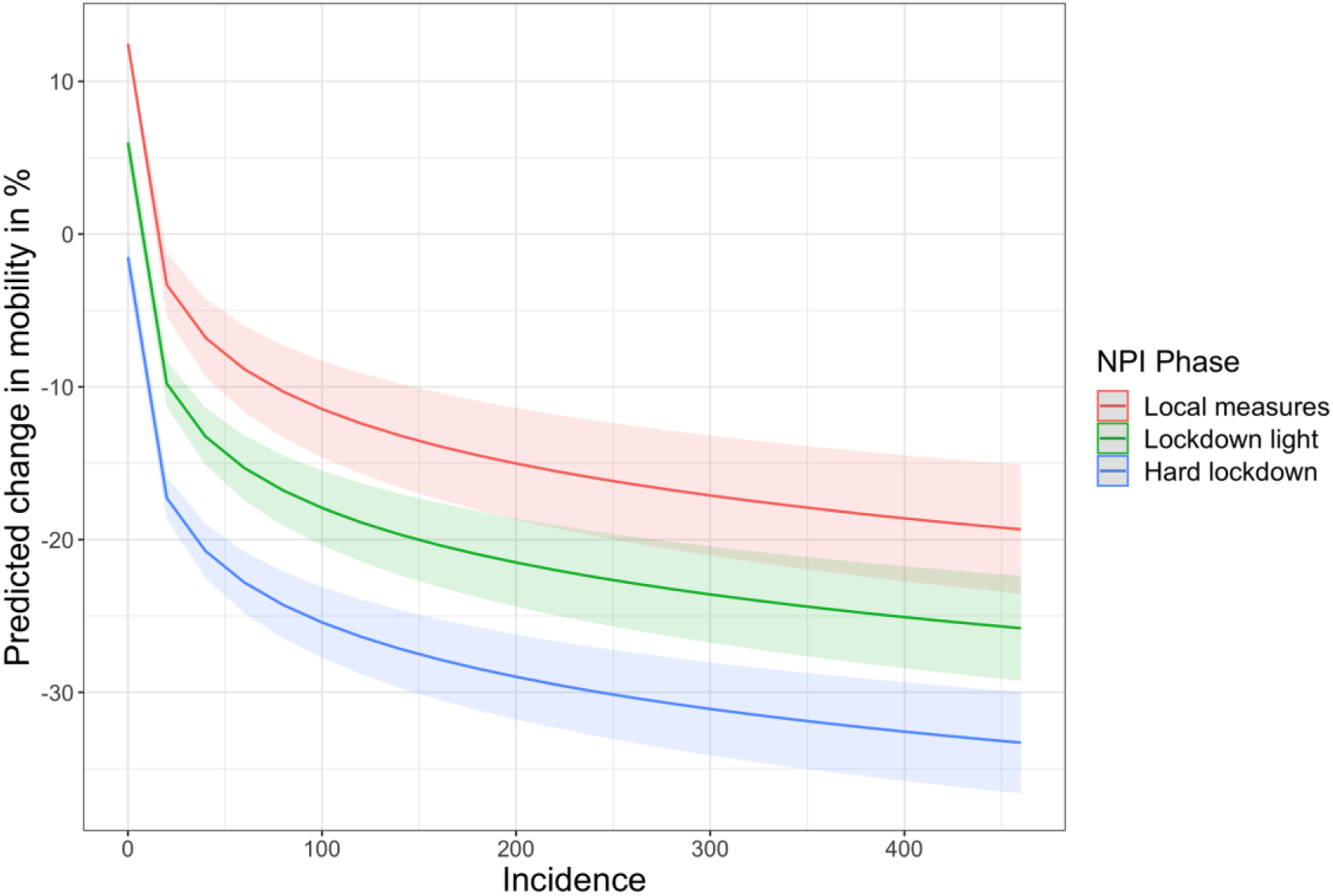
Marginal effect of incidence on mobility during three phases of NPI deployment. The plot visualizes the marginal effect of 7-day incidence on predicted relative changes in mobility (compared to 2019 baseline) during three successive phases of national NPI response: NPIs implemented at county-level (until Nov 1), a national-level “lockdown light” (until December 15) and a full-scale national lockdown (from December 16). The plot is based on Model D presented in S1 and was generated using the R package *ggeffects* [99].

#### 4.1.3 Risk signals & public attention: A comparative analysis

While these results suggest a significant and pronounced behavioral adaptation during the “second wave”, the overall behavioral response was less pronounced than during March of 2020. In the top panel of Fig 5, mobility and incidence data are juxtaposed for the early pandemic (March 2020) and the second wave (Oct 20-Jan 2021). The data indicate that in spite of higher disease prevalence, the overall reduction in mobility was lower. In the following analysis, we contextualize this with a brief comparative analysis of data available at the national level for Germany, focusing on risk signals emanating from political decisions and public attention. For this, we consider data from traditional and social media as well as from the *Google* search engine as an indicator for information-seeking behavior (Fig 5).

**Fig 5.**
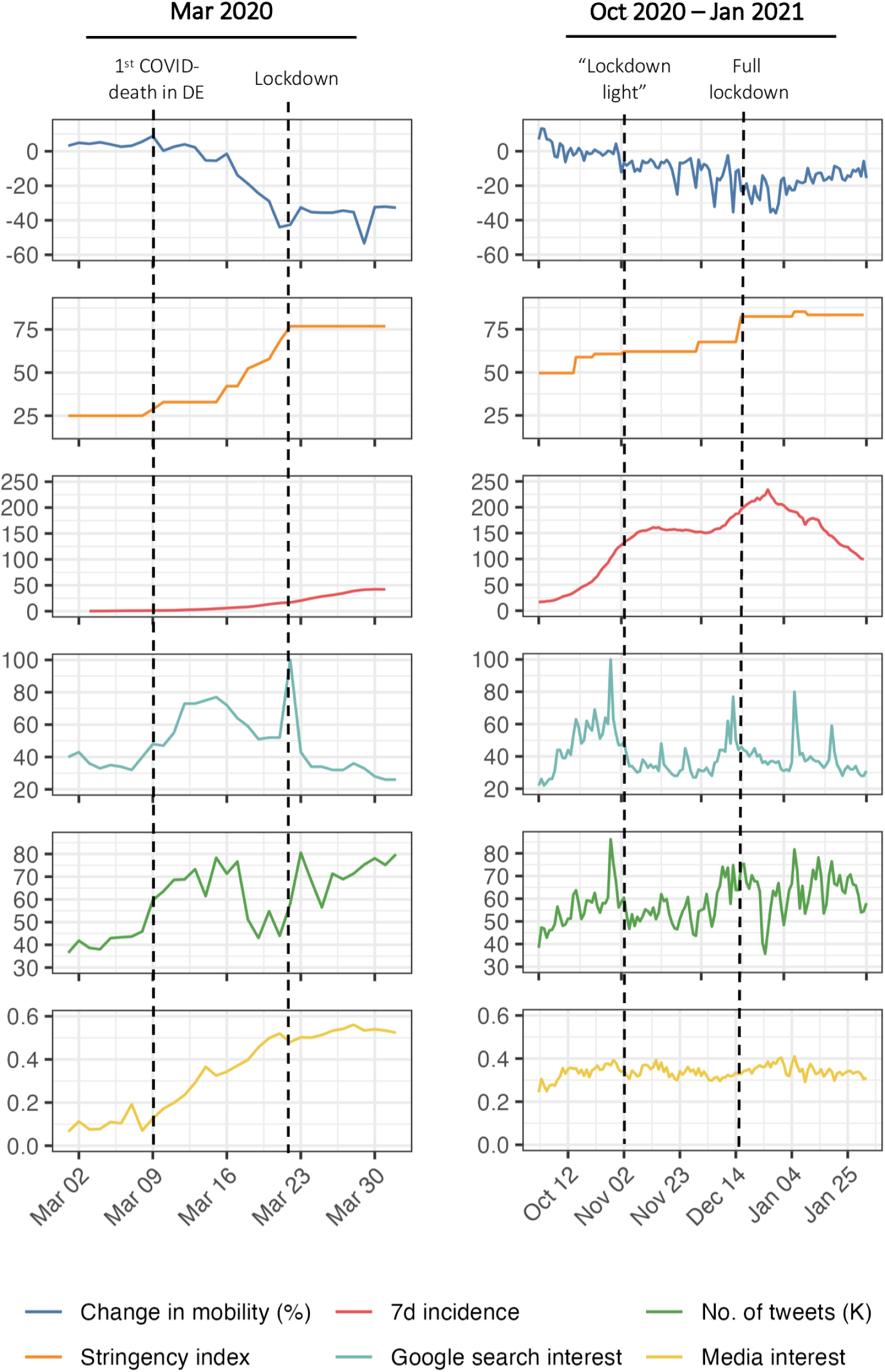
Pandemic dynamics and public visibility in Germany. The plot depicts various data [39, 98, 103–105] related to pandemic dynamics and public interest for Mar 2020 (left side) and Oct 2020 – Jan 2021. Note: *Media interest* refers to the share of articles in 68 national German online news media that mention “Coronavirus” or “COVID-19”. *Google search interest* is an index for the search interest in a topic over a specified period of time ranging from 0-100.

The early pandemic in Germany was, arguably, a period characterized by clear risk signals and high public attention. Building on a broad consensus between political decision-makers and scientists, NPIs were gradually ramped up, e.g., by restricting large events, culminating in a national lockdown by March 22. As the left side of Fig 5 indicates, private mobility had already fallen drastically within the two weeks prior to this first lockdown, indicating a significant autonomous response. This, however, occurred in lockstep with the stepwise announcement and enactment of increasingly stringent containment measures. Thus, it seems likely that individuals estimated the benefits of adaptation (i.e., avoided risks) to be very high against the background of limited information on the virus, high levels of fear and high public awareness. This interpretation is reflected in the data in the bottom three panels of Fig 5, which indicate a substantial increase in interest of both traditional and social media for COVID-19, along with an increase in private information-seeking. As has been argued by others [100], the degree of political determination to curb the spread of the virus resulted in heightened awareness and support for response policies, which enabled the success of Germany’s initial response. Or, in other words, a clear risk signal emerged from the policy response in March 2020, which may have contributed to an anticipatory autonomous response.

In the fall of 2020, the situation was less clear-cut. A more controversial debate as to which course should be taken had emerged: The different actors involved in decision-making in the federal system in Germany required longer to agree on a coordinated response, which was less decisive than during the first wave [101]. As we described above, this resulted in heterogeneous local response measures followed by the “lockdown light”. This ambiguity seems to be reflected in the data on public attention: The bottom three panels of Fig 5 indicate that despite continuously high infection levels in November, the attention placed on the pandemic in (social) media and in Google searches remained comparatively low, while only events such as the initiation of national lockdowns are accompanied by attention spikes. As we address in more detail below, this period also roughly coincides with a marked decline in trust in the government and the first large-scale anti-containment demonstrations [102]. The lack of cohesion with respect to the severity of the situation and the adequate response is reflected in the rather constant level of mobility in top right panel of Fig 5. After hospitalization levels reached critical levels [101] and the national lockdown was initiated mid-December, a reduction in both mobility and case numbers was achieved in a comparatively short amount of time.

### 4.2 Time variance of behavioral adaptation

The comparison between spring and fall of 2020 underscores that the relationship between autonomous and policy-induced adaptation is unlikely to be constant over time. As we explored in Section 3.3, this may be the case because key determinants such as risk perceptions or trust in government change over time. Here, we investigate whether this has been the case in Germany, using the most detailed publicly available data set dealing with attitudes toward COVID-19 the authors are aware of [106]. The data stems from a representative longitudinal survey of the general public in Germany, conducted in 48 waves with an average sample size of around 1,500 interviews. Detailed information about the data as well as methods used in this section are presented in Supplementary Information S2.

#### 4.2.1 Diminishing risk perception

It is likely that preventative behaviors decrease if the perception of risks associated with an infection declines over time. To investigate whether this has been the case in Germany, we statistically analyze the relationship between incidence, as a measure of the current epidemiological risk, and risk perception. As we explain in more detail in S2, the data by [106] includes three questions relevant for risk perceptions which were asked at irregular intervals in 29 survey waves. However, due to some constraints in matching the perceived risk to state-level incidence data [98] we can only make use of data from 21 waves. We construct a simple composite risk perception variable from the three relevant variables and calculate state-level averages. This results in a sample size of 21 pairs of incidence and risk perception data for the 16 German states between August 2020 and April 2022 (n = 336). As a base model, we establish the association between perceived risk (response) and incidence as well as a numeric variable measuring the days since the first observation (predictors) in a simple linear model. Due to their ability to deal with small sample sizes and unevenly spaced time series data, we then fit the data to linear mixed-effect models [107, 108], including state-level and date-level random effects. In S2 Table 1 we provide detailed regression results and discuss model diagnostics. Across all models, the results indicate a consistent and significant relationship between incidence (logarithmic form) and average perceived risk. The linear variable measuring the progression of time shows a negative sign and high significance. The addition of state-level and date-level random effects significantly improves model fit, indicating that some unobserved heterogeneity and seasonal effects can be captured through this. In sum, the results suggest that at a given level of incidence, the average degree of perceived risk declines over time. In Fig 6, we use our model with the best fit to visualize this effect: Holding incidence constant at the median value of our data set, the predicted perceived risk decreases over time. While these outcomes can merely be seen as tentative due to small sample size and geographic aggregation, they seem to substantiate the idea that infection levels become less intimidating as time passes. Our analysis cannot, however, provide an explanation of why this occurs, as several complex issues may interact, such as a better ability to manage risks through testing and vaccinations or an overall habituation to infection risk. Nonetheless, we can infer that a relevant predictor of behavior has changed significantly over time, which may impact both autonomous adaptation and compliance rates.

**Fig 6.**
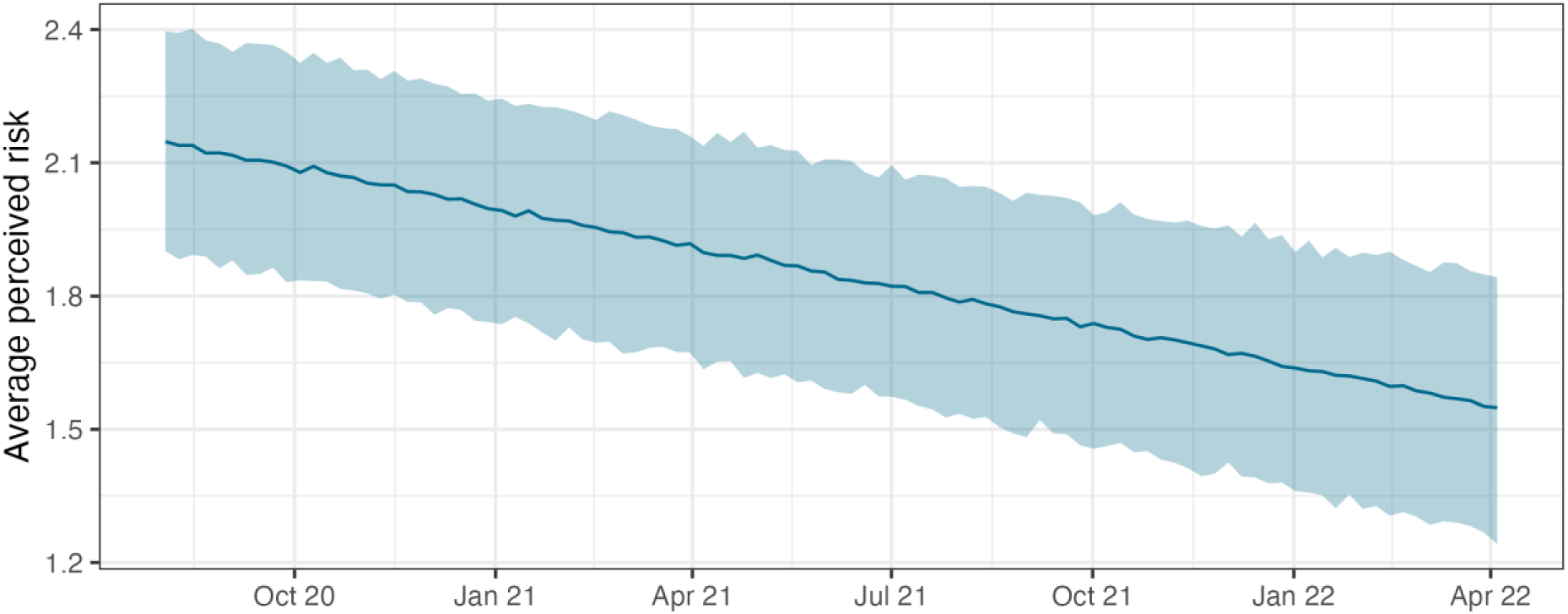
Diminishing risk perception over time. The plot depicts the marginal effect of time (measured as a linear variable) on perceived risk, beginning in August 2020 and assuming a constant level for 7-day incidence. The figure was generated using the coefficients of Model C (cf. Supplementary Information S2) and the median incidence value in the sample (105.84). The light blue ribbon indicates a 95% prediction interval, generated with the R package *merTools* [109].

#### 4.2.2 Erosion of trust and compliance

Longitudinal studies indicate a marked decline in trust in the government and its ability to manage the pandemic in Germany, starting in the fall of 2020: While in a bi-weekly survey, 65% of respondents thought the government handled the pandemic “somewhat” or “very” well on October 7, 2020, this number decreased to a mere 21% by March 24, 2021 [110]. As trust is often considered a key factor for behavioral adaptation, its development over time may provide significant insights for compliance with NPIs (see Section 3.2). To investigate a potentially decreasing degree of compliance and its association with trust, we analyze once more data collected by [106], focusing on two recurrently asked questions: (i) the degree to which respondents perceive information provided by the government about the COVID-19 pandemic to be credible and (ii) whether they perceive the measures taken by the government as adequate, insufficient or excessive. For these questions, data in ordinal response categories is available from 36 survey waves (n= 43,106, for details see S2). Fig 7 plots the data over the observed time span from April 2020 to April 2022 and indicates a fluctuating, yet overall declining share of respondents considering government information to be credible and those believing that containment measures are adequate.

**Fig 7.**
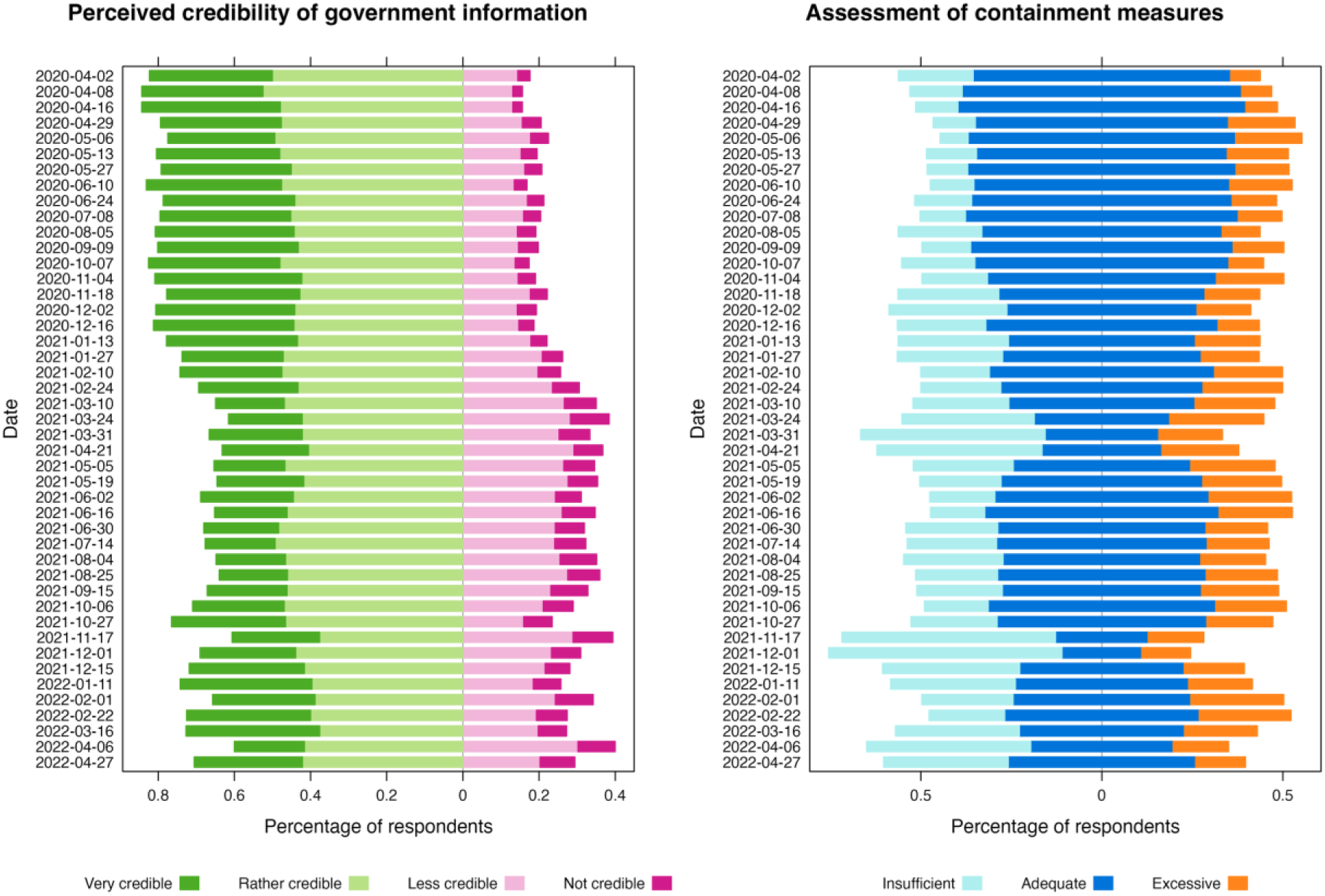
Credibility of government information and assessment of containment measures. Assessment of the credibility of information issued by the German government about COVID-19 (*left*) and assessment of the adequacy of containment measures (*right*). Data: [106]

Assuming that the perception of containment measures is directly or indirectly related to the degree of compliance [55], the relationship between trust and agreement with measures may be interpreted as an, albeit imperfect, proxy for the relationship between trust and compliance. In Fig 8, the association of both variables is illustrated by two plots: The ‘mosaic plot’ on the left-hand side provides a visual description of how both variables are related. On the right-hand side, a conditional effects plot [111] depicts the results of an ordinal regression model using perceived credibility of information provided by the government to predict agreement with measures. As the plot indicates and we discuss in more detail in S2, the model robustly relates the assessment of measures with the respondent’s perception of credibility of government information. With decreasing perceived trust, the probability for respondents to consider measures excessive increases considerably. Conversely, high perceived credibility is associated with a higher probability of finding that measures do “not go far enough”. This, along with the data presented in Fig 7 may be interpreted as an increasing fragmentation of public opinion, indicating that an increasing share of the population distrusted governmental information and believed measures to be excessive, which may likely explain observed increases in non-compliance. On the other hand, as the light blue lines in Fig 7 indicate, an increasing share of respondents also considered measures insufficient, which may suggest higher autonomous efforts for infection prevention. While others have treated compliance in more detail [112], these findings further substantiate that significant changes occurred over time in one of the key determinants of behavioral adaptation.

**Fig 8.**
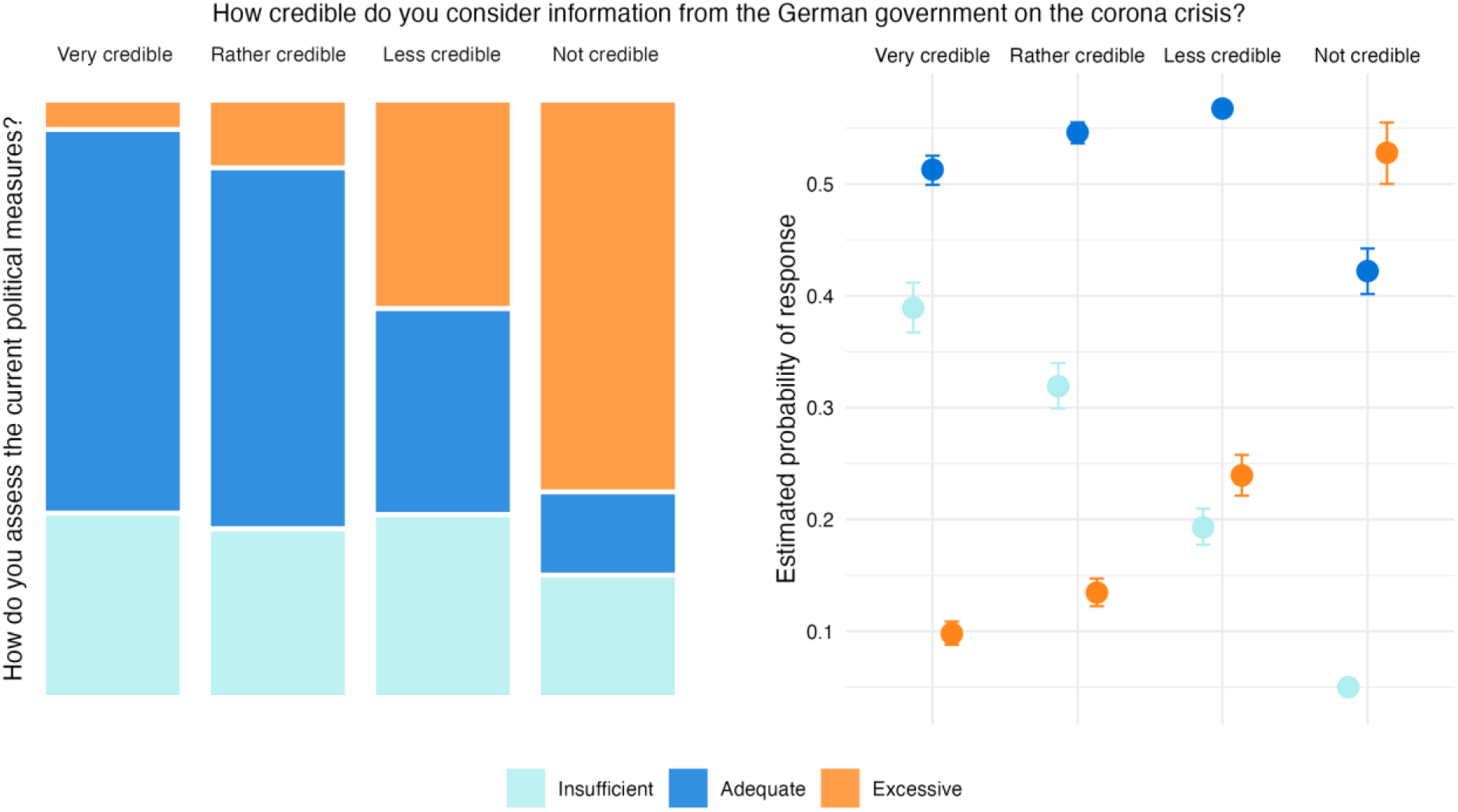
Association of perceived credibility of government information with assessment of containment measures. Left side: Each segment indicates a specific combination of response categories in the data set. Right side: Conditional effect of perceived credibility of information from the government on assessment of containment measures. The posterior mean estimate of the probability of responses in each opinion category is shown for each of the four categories of perceived credibility, with error bars indicating 95% credible intervals.

## 5. Modeling autonomous and policy-induced adaptation

While the primary focus of this analysis is to better understand the interplay of autonomous and policy-induced adaptation conceptually and empirically, both are directly related to behavioral-epidemiological modelling. Thus, in this Section, we briefly discuss the relevance of this analysis for modeling efforts, focusing on two key areas:

1) *Assessing the representation of behavioral adaptation in existing modeling frameworks*: Building on a non-exhaustive inventory of modeling literature, we use our framework to give an overview on how the previously discussed determinants and interactions are represented in existing models. Based on this, we discuss promising avenues for future model developments (Section 5.1).
2) *Developing conceptual models to understand system dynamics*: We discuss why an improved understanding of behavioral adaptation is a prerequisite for developing counterfactual behavioral responses in simulation models. We showcase how conceptual models may provide useful insights on the interplay of autonomous and policy-induced adaptation in different situations (Section 5.2).

### 5.1 Representation of autonomous and policy induced adaptation in behavioral-epidemiological models

The following is intended as a brief overview of how autonomous and policy-induced adaptation processes have been modeled, to then discuss to which extent their effects can be disentangled and changes over time can be represented. It is not an exhaustive treatment on how human behavior can be represented in epidemiological models [22, 113, 114] nor a review of specific models of the COVID-19 pandemic [115, 116], as both are beyond the scope of this paper. To structure the overview, we focus on two common types of mathematical models namely (i) agent-based models and (ii) models based on differential equations (cf. *Note 5*). In agent-based models (ABMs), populations are represented by agents endowed with specific rules on how to interact in a given environment. In the context of the COVID-19 pandemic there are vast applications and different meta studies with varying time spans and focus points [e.g., 115, 117]. One of the strengths of the ABM approach is that it allows the representation of agents at the micro level and the incorporation of heterogeneity with respect to socio-demographic characteristics, location, specific behavioral patterns and more. Differential equation models (DEMs) reflect infection dynamics at the macro-level using different population compartments and are computationally less expensive, faster to develop and more readily interpretable. Here, likewise, numerous applications and reviews have been contributed [e.g., 118, 119]. In both ABMs and DEMs, relevant human behaviors for transmission are typically physical contacts and mobility. DEMs are usually based on the assumption of random mixing in the population or subsets thereof [120], with transmission between the different compartments occurring at a specific probability. While the majority of ABMs also relies on such assumptions, according to a review [115], some ABMs assume more realistic, agent-specific contact behavior. These can be based on real-world contact networks or activity patterns [121, 122] and take into account relevant factors such as the degree of infectiousness of specific agents, wearing of masks, duration of contact, air exchange and more [123, 124].

In assessing behavioral adaptation, the vast majority of modeling studies seems to focus on policy-induced adaptation, i.e., the effect of individual or multiple NPIs on contacts or mobility. In models based on random mixing, policy-induced adaptation is represented as a reduction of the number of possible contacts or a reduction of the transmissibility of given contacts, for instance assuming effects of masks or vaccination [123]. In agent-based models, agents may change their specific contact patterns as a result of testing or contact tracing [123, 125, 126], after they themselves or others have become infected [127, 128] allowing them to subsequently only encounter a subset of their contacts. Compliance with NPIs has been introduced in some models, for instance by splitting the population in different compartments and estimating parameters for compliers and non-compliers separately [129].

Autonomous adaptation, on the other hand, is included less frequently in models. In DEMs, autonomous adaptation processes have been introduced by endogenizing a response in the contact rate to certain state variables, usually the number of infected or dead [130], assuming that these signal risk to the individual. This is for instance accomplished by the introduction of a new compartment representing risk perception [131] or by setting contact rates through a utility maximization process, in which social contacts increase utility and the risk of an infection decreases it [24]. Such models frequently result in other long-run dynamics than those without autonomous adaptation, by finding an equilibrium at a reproduction number of 1. In ABMs, adaptive behaviors have been represented in higher detail, for example when agents decrease contacts in proportion to the number of cases in their area [126] or their network of personal contacts [125]. However, despite their ability to incorporate heterogenous behaviors, merely about 5% of ABMs included adaptive behaviors in the review by [115].

Fig 9 summarizes this brief overview. In light of our analysis, approaches that address how the behavioral response changes over time are of particular interest. Time-varying parameters are widely used in behavioral-epidemiological models, for instance a time-dependent contact rate, which may be obtained by using proxy data from mobility data sets [132, 133] or by directly fitting models to the data [16, 134]. Such approaches have been successful in reproducing observed case numbers and death rates. However, neither approach allows a direct interpretation as to *why* contact patterns have changed because that process is not endogenous to the model. As we expound further below, however, it is of key interest whether a change in behavior is the result of policy, of autonomous adaptation or both.

**Fig 9.**
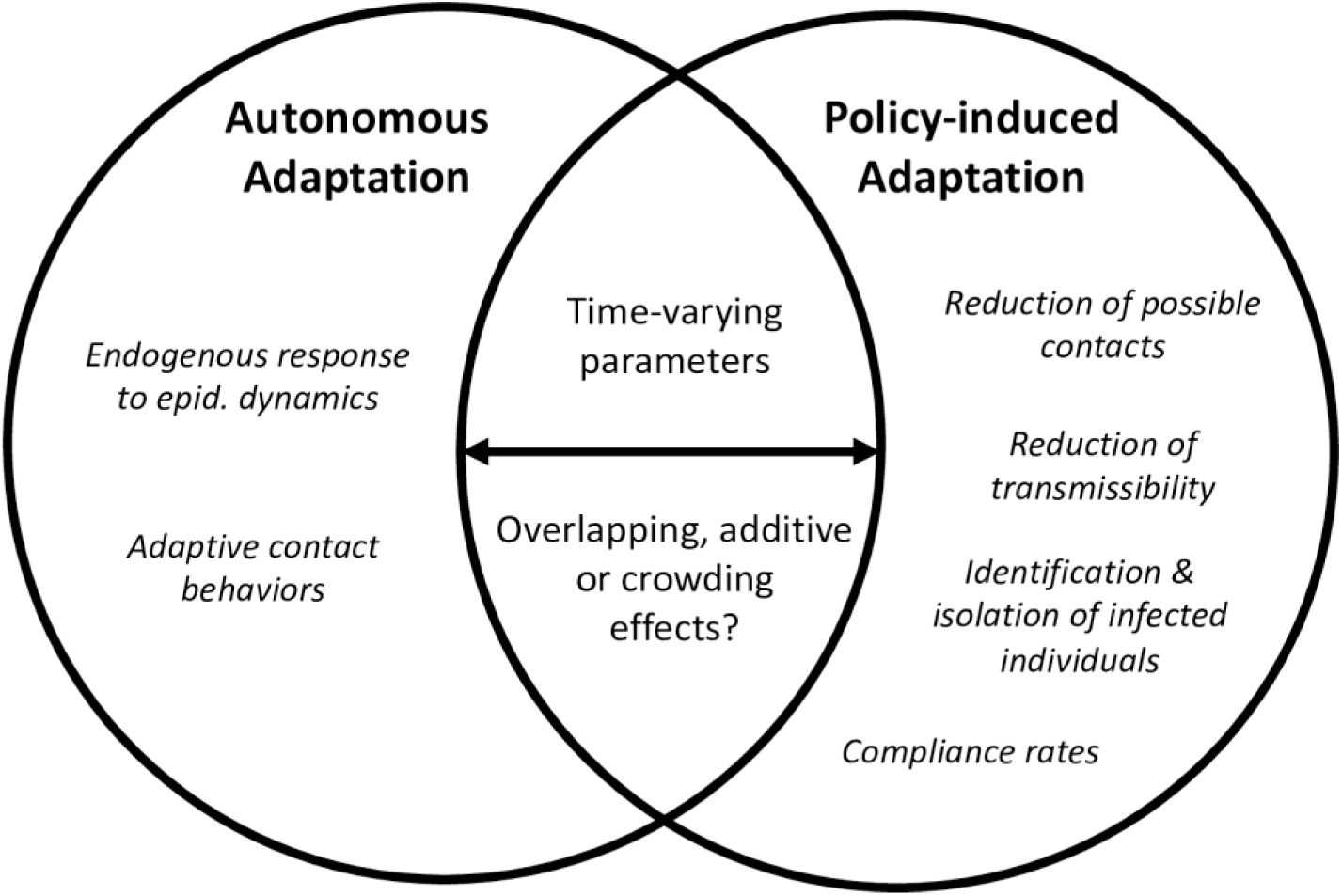
Confluence of autonomous and policy-induced adaptation in behavioral-epidemiological models. In behavioral-epidemiological models, changes in contact patterns and mobility result are modeled through varying mechanisms related to autonomous and policy-induced adaptation. However, how the two adaptation mechanisms come together (“confluence”) has not been addressed sufficiently.

Thus, it is promising that an increasing number of models includes the effects of both autonomous and policy-induced behavioral adaptation [11, 16, 135–137]. Here, the key question is how the two effects are set in relation to one another. Do they overlap, add to one another, or is there a crowding effect? In [135], for example, NPIs set the boundary conditions for the number of possible contacts, which then vary in dependence of ICU occupancy. [137] characterize their relationship as additive and estimate the parameters for both effects concurrently from data. In their approach, identifiability is related to the dynamics of incidence number and change points of policy response. When the incidence number is rising (or falling) rapidly, the complementary or interactive effects of both autonomous and policy-induced adaptation should be detectable [138, 139]. If, however, the incidence is in a plateau, and the deployment of NPIs changes, identifiability issues may arise and a separate estimation of the two parameters may be impossible. [11] circumvent such issues by combining an econometric analysis of mobility with a ‘controlled SIR’ [140] which allows them to decompose the changes in the contact data inferred by the model into separate effects for autonomous and policy-induced adaptation.

### 5.2 Exploring system dynamics and policy strategies through conceptual models

Disentangling the effects of autonomous and policy-induced adaptation and understanding how they interact is highly relevant for assessments of the effectiveness and cost of intervention strategies. The support of policy decisions can be considered the primary use for behavioral-epidemiological models [141], which require adequate assumptions for counterfactual scenarios. As [136] point out, any narrative such as “strategy X would have saved more lives” is built on (implicit) behavioral assumptions. If that assumption is that no autonomous adaptation occurs, the counterfactual to compare interventions against may assume substantial exponential growth of infections [e.g., 142, 143] and thus overestimate the role of interventions based on observed data [8, 9]. Similarly, both behavioral adaptation processes have to be considered when evaluating the cost associated with a political response. Lockdowns, for instance, have been considered costly due to a slowdown of economic activities. If a substantial part of mobility reduction, however, is driven by autonomous decisions [6] then the ensuing macroeconomic cost cannot be attributed to the lockdown alone [11].

Conceptual models, simulating behavioral adaptation under different assumptions, may help to better understand system dynamics, particularly the interplay of autonomous and policy-induced adaptation, and thus provide a basis for sound counterfactuals. During the pandemic, data-driven forecasting models were of key importance to provide immediate decision support. The use of behavioral theories and parameters in infectious disease models, however, has been characterized as inconsistent by review analyses [1, 21], not least because of lacking data (e.g., on risk perceptions) and the complexity of the interplay of behavioral or social factors. Thus, an analysis of adaptation mechanisms and their interplay in a conceptual way may improve our grasp of system dynamics and policy impacts under a range of different assumptions and scenarios. A number of conceptual and policy-simulation models has been developed, analyzing different timings and duration for interventions [143], impacts of a combined autonomous and governmental response [144], or detailed systems models including contact behaviors and economic processes [145].

However, we are not aware of systematic analyses characterizing the interplay of autonomous and behavioral adaptation under different assumptions. While an extensive treatment of this is beyond the scope of this article, we underscore how this could be useful by adapting a simple SIR model [146]. As we present in detail in Supplementary information S3, we assume a small population with a time-varying contact rate. This contact rate is adapted in response to containment policies, i.e., NPIs result in a direct reduction of contacts by a specific percentage, implemented as a smoothed jump. We use an existing specification from the early pandemic [24] to represent an autonomous response based on an expected utility framework, where the contact rate is reduced in response to the number of infected. For simplicity, we assume no interaction between both adaptation mechanisms but let their effects overlap. Note, however, that this is a simplifying assumption for the sake of illustration and should be relaxed by later, more in-depth analyses. Consider the example presented in Fig 10: We compare an early intervention after 7 days (left-hand panels) to a later response (right-hand panels, after 21 days). The bottom panels indicate how the assumed impacts of autonomous and policy-induced adaptation affect the contact rate: In the case of the early interventions, behavioral adaptation is driven mainly by policy. In the case of the later intervention, the initial increase in infections results in significant autonomous adaptation driving the early behavioral response, whereas the effect of policy only sets in later.

**Fig 10.**
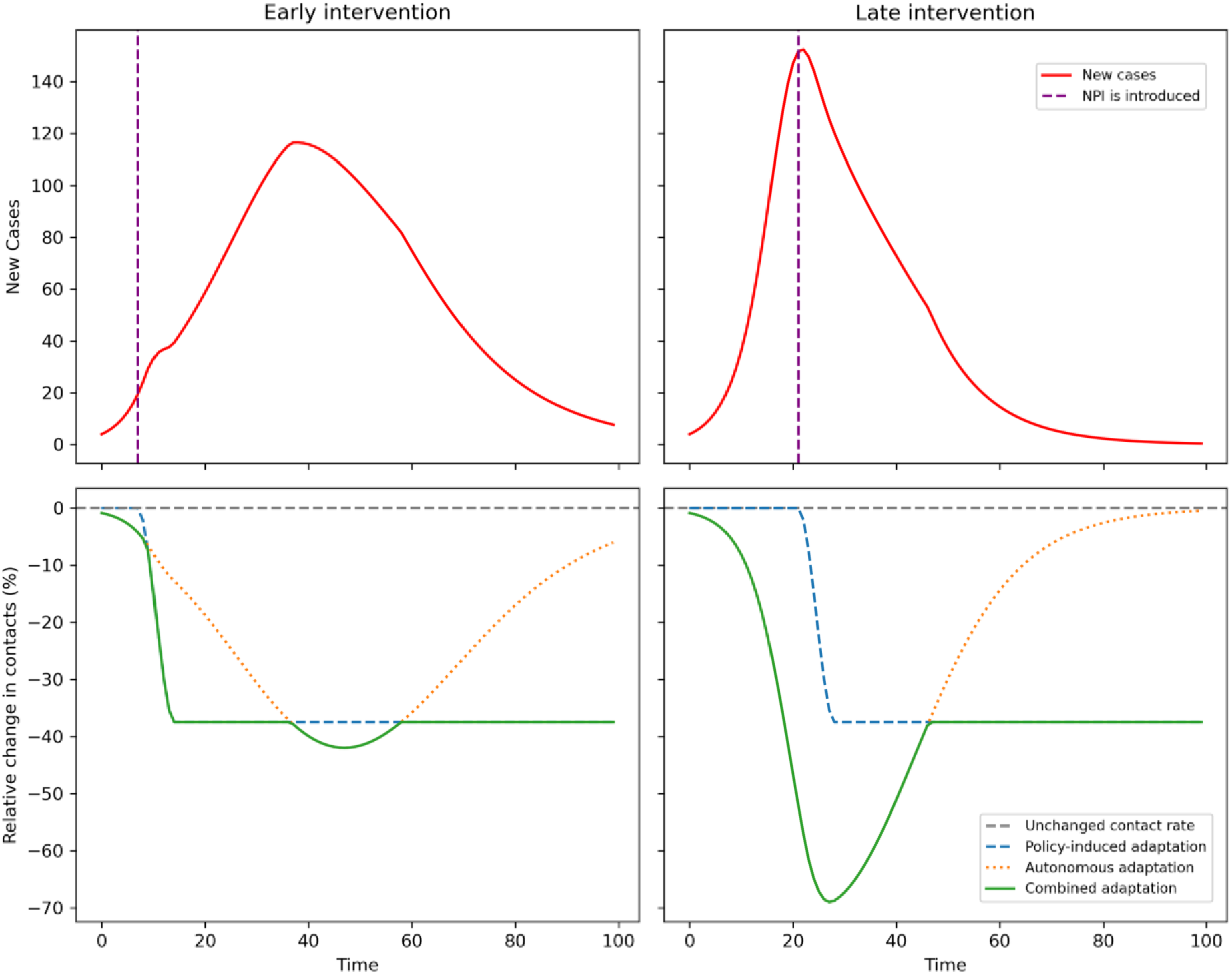
Autonomous and policy-induced adaptation, contact rates and infections. The top panels show the number of daily new infections for an “early” and “late” intervention (dotted line, after 7 and 21 days, respectively). The bottom panels show relative changes in contact rates resulting from policy (blue dotted line), autonomous response (orange dotted line) and the combined effect (solid green line). Details on model specification can be found in Supplementary information S3.

Similar to understanding the interplay, such models could be used to gain perspective on the sensitivity of models to certain parameters. In Fig 11, we exemplify this by running our conceptual model under three different assumptions for the risk aversion parameter, which regulates the strength of the autonomous response to the number of cases (see S3). Assuming an intervention after 21 days, the model shows distinct differences between very risk averse and risk tolerant populations. Such analyses may allow differentiating the impact of certain intervention strategies under heterogenous risk preferences. Thus, well established differences in risk perceptions within or between populations [35, 147] may be considered and represented.

**Fig 11.**
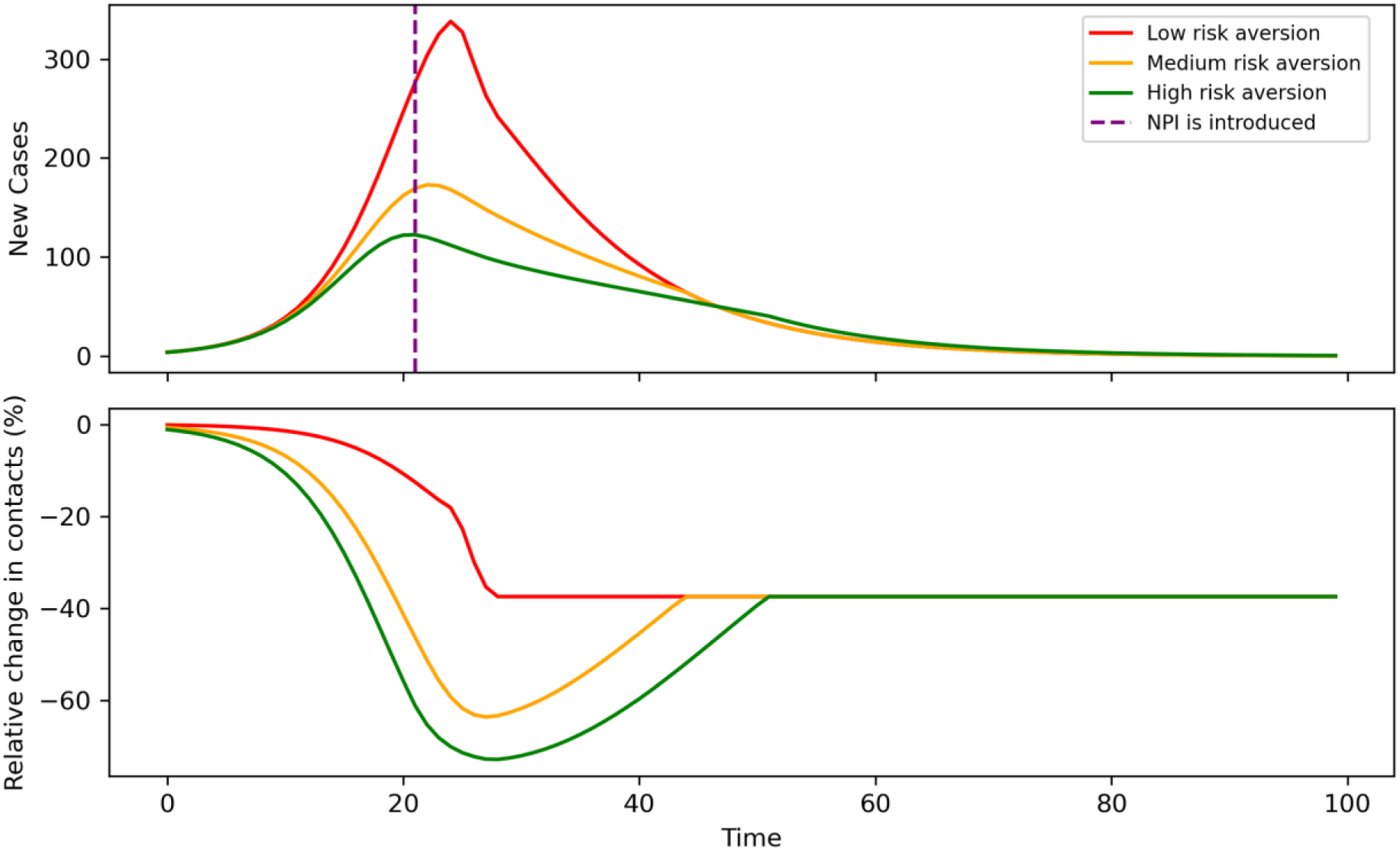
Exemplifying the impact of different degrees of risk aversion. The top panel shows the number of daily new infections under three different assumptions for risk aversion in the population and the date of intervention (vertical dotted line). Bottom panel: Corresponding relative changes in the contact rate under the three assumptions. For details see S3.

Even though these are deterministic examples with deliberately set effect sizes, they can showcase which type of counterfactual analyses of autonomous and policy-induced adaptation may be carried out with conceptual models. Later, more in-depth analyses could characterize interactions between both adaptation mechanisms (e.g., impact of risk signal from NPI introduction to autonomous response) or the impact of time variance on key parameters (e.g., declining contact rate or risk perception) in higher detail. When the mechanisms driving behavioral adaptation or the change in contact rate are understood better, different response strategies may be evaluated more accurately with respect to their effectiveness at disease prevention and their costs [136].

## 6. Discussion

Understanding how human behaviors influence infectious disease transmission is essential. This article contributed towards analyzing and modelling explicitly two interrelated forms of behavioral change in a pandemic: Autonomous and policy-induced adaptation. Synthesizing insights from various strands of behavioral literature on COVID-19, we characterized both adaptation mechanisms and their determinants. Autonomous adaptation refers to voluntary behavior changes due to a number of socio-demographic and personal factors, particularly perceptions about infection risk and efficacy of preventative actions [29, 31, 34]. Policy-induced adaptation occurs when individuals alter their behavior in response to non-pharmaceutical interventions and is influenced by factors such as compliance levels, social norms, and implementation strategies [46, 57, 59]. Though both forms of adaptation are distinct, their predictors overlap and they can be difficult to disentangle empirically. To address this challenge, we developed an analytical framework focusing on confluence of autonomous and policy-induced adaptation, their interactions and changes over time. This framework understands behavioral adaptation as a “moving target”, where autonomous and policy-induced adaptation overlap and diverge in a dynamically evolving interplay. The two adaptation mechanisms interact, which was analyzed with a focus on risk signals and trust: Among others, government activities can indicate risk and thus influence voluntary behavioral changes [10, 17]. Trust in government, science and media, on the other hand, will affect whether such signals are heeded and to which extent individuals comply with behavioral mandates [84]. This interplay evolves over time, e.g., when effects such as a diminishing risk perception or an erosion of trust set in, potentially altering the relationship and relative importance of both adaptation mechanisms. Due to its flexibility, this framework may provide a useful system of reference for future research. Here, it formed the basis for our analysis of the German case and for the subsequent discussion of how autonomous and policy-induced adaptation can be represented in behavioral-epidemiological models.

Our empirical investigation of Germany focused on the fall of 2020 and analyzed changes in mobility in response to predictors such as the stringency of policy response and 7-day-incidence as proxy for infection risk. The statistical analyses indicate that both autonomous and policy-induced adaptation resulted in relevant reductions of mobility during the second wave of the pandemic. This is in line with findings from the early pandemic [8], albeit with an overall weaker effect than in March of 2020. Our analysis of risk signals during the first and second wave demonstrated significant differences in the decisiveness of policy response [101] and public attention for the pandemic. This substantiates that the perceived severity of the situation may have been different, at least for parts of the population. Relying on data from a large survey panel [106], we also examined changes over time in key determinants of behavioral adaptation, focusing on diminishing risk perceptions and the erosion of trust. Our analysis of risk perceptions provided tentative evidence for a declining association between infection levels and the subjective impression of risk over time, which others have reported as well [50]. It is important to note, however, that the analysis was carried out using a comparatively small sample due aggregation of data. Potential explanations for a changing assessment of infection risk include increasing knowledge about the virus and the associated risk, habituation effects and increasing access to vaccines as well as other methods to mitigate infection risk (e.g., rapid antigen tests). In a second analysis based on the time series, we found that the German government lost credibility in the eyes of a substantial part of the population. According to our ordinal regression analysis, those distrustful of government information were substantially more likely to view containment measures as excessive. Given the linkage established by others between feeling ‘disinformed’ and non-compliance with NPIs [148], this provides an indication for eroding compliance in parts of the German population. In sum, these analyses provide several sound indications that autonomous and policy-induced adaptation are subject to significant changes over time.

To accurately represent complex behavioral adaptation processes – be they driven by policy or self-protection – in parsimonious models is highly challenging. This is particularly so as reliable data on perceptions and attitudes may not be available in high resolution or real time. This explains why many modelers adopt pragmatic, data-driven approaches and the use of theories of behavior change in infectious disease modeling has been characterized as “patchy” [20]. Notwithstanding, particularly autonomous adaptation processes should receive more attention in infectious disease models, as others have argued [149]. Based on our literature and empirical analysis, we discussed relevant avenues in behavioral-epidemiological modeling: Emerging approaches combine endogenous autonomous adaptation with representations of policy impacts, thus including both autonomous and policy-induced adaptation [135, 137]. Future work should increasingly focus on their interplay and why parameters vary over time. Beyond reproducing disease outbreaks accurately, it is relevant to know the reasons driving contact rate changes, which may be accomplished by either endogenizing the mechanisms or providing contextual analyses. Such advancements would be of high value for the scenario-based analysis of intervention strategies. Behavioral adaptation needs to be understood to construct convincing counterfactuals and analyze the effects of policy interventions on behavior. As we argued in the previous section, conceptual and policy-simulation models may help to gauge system responses under various assumptions. Merely considering the deployment of NPIs ignores relevant aspects of how behavior shapes infectious disease transmission. Only if both autonomous and policy-induced adaptation are accounted for, can the impact of interventions on public health be adequately determined [8, 9] and the associated cost better understood [11]. However, it is also important to note that beyond medical cost and the decrease of economic activity, behavioral mandates impose cost on society for instance through increased incidence of mental health issues and domestic abuse, increases in preventable deaths, education deficits, and restriction of civil liberties [150, 151]. Thus, it is also of high relevance to know to which extent self-protection efforts can replace mandates [152] as voluntary action tends to be less costly [153]. It is also relevant to note that approaches perceived to be controlling can result in a loss of trust, for which our analysis in Section 4.2 provides some indication.

This article is limited by a number of constraints. We approached a complex topic with many nuances, which implies that omissions and emphases cannot be avoided. For one, in our perspective on autonomous and policy-induced adaptation, social norms and processes have only been touched upon lightly, whereas they likely carry significant weight [50, 51]. Moreover, a variety of contextual conditions are highly relevant for behavioral adaptation, including factors such as political culture and other national framework conditions, which we did not address in higher detail. Our empirical analyses were limited to publicly available data sets, resulting in issues matching data from different sources and the need for geographic aggregation. In our analysis of autonomous and policy-induced adaptation, for instance, we found the German federal states to be the smallest shared geographical unit. However, some of the German states are rather large and have distinct regional heterogeneities (e.g., rural vs. urban), for which a more fine-grained analysis would have been beneficial. Nonetheless, our results are in line with other analyses based on data in higher spatial resolution, indicating that key effects can be found with a comparatively parsimonious approach. Further disaggregation would have particularly benefitted our analysis of diminishing risk perceptions, which was based on a small sample. It is thus important that future work revisits and corroborates these findings.

## 7. Conclusion

This article contributed to the emerging understanding, analysis and modeling of autonomous and policy-induced behavioral adaptation during a pandemic, with a specific focus on their relationship and development over time. We developed a more precise behavioral framework by synthesizing insights from various bodies of literature on behavior change during the COVID-19 pandemic, focusing on key determinants (e.g., risk perceptions) and interactions (e.g., risk signals & trust) between autonomous and policy-induced adaptation. Both mechanisms and their relationship likely evolve over time, for instance when individuals become desensitized to infection risk or develop aversion against NPIs perceived to be controlling. We applied the framework in an empirical analysis of the German case which demonstrated that during the “second wave” of COVID-19 in the fall of 2020, mobility patterns changed significantly due to both autonomous risk management and containment measures. However, mobility reductions were smaller than in the early pandemic, which may be explained by ambiguous risk signals and lower public attention. Through analysis of survey data we found indications that there is a diminishing relationship between infection levels and risk perceptions, and that a substantial share of the population lost trust in information provided by the German government. Both trends likely further reduced the use of preventive behaviors over time. Against this background, a brief discussion of the representation of behavioral adaptation in epidemiological models was carried out. While promising modeling approaches are developed, it is key to further disentangle the effects of autonomous and policy-induced adaptation and accurately represent their interplay. Conceptual models may improve our understanding of how both effects interact and evolve and therefore support the development of counterfactual scenarios. By doing so, the impacts of alternative intervention strategies can be evaluated in a more convincing way, with high relevance for future pandemic management.

## Data Availability

All relevant data for this manuscript are publicly available. References are made throughout the manuscript and in the Supporting information. The specific data used in Section 4 can be accessed here: Incidence data is available from https://zenodo.org/records/8267744. Data from longitudinal study of COVID-19 perceptions in Germany is available from: https://doi.org/10.4232/1.14032. Data on state-level stringency is available from https://www.corona-daten-deutschland.de/dataset/massnahmenindex_bundeslaender_pro_tag. Data on changes in mobility is available from https://www.destatis.de/DE/Service/EXSTAT/Datensaetze/mobilitaetsindikatoren-mobilfunkdaten.html. Further data plotted in Fig 5 is available from https://www.mediacloud.org/, https://trends.google.com/trends/, https://crisisnlp.qcri.org/tbcov

https://trends.google.com/trends/

https://www.mediacloud.org/

## Notes

### Note 1

The effect of human behaviors on the spread of a contagion can be further differentiated than we do here. There are, for instance, relevant differences between reducing contacts or adopting measures that reduce the probability of transmission of physical contacts (e.g., use of facial masks). Here, due to our focus on behavioral change and its drivers, we do not differentiate types of behavior for simplicity.

### Note 2

Non-pharmaceutical interventions may have a variety of indirect effects on behaviors. The retention of reserve beds in hospitals, for example, may incite some individuals to take higher risk assuming that they can be treated. Here, however, we focus on more direct policy impacts for simplicity.

### Note 3

Note that misinformation has often played a critical role here, with interactions to the social media sphere [93].

### Note 4

Trust in government can become a double-edged sword, however, as a case study of Singapore showed: If the competence of the government is believed to be high, individuals may reduce their own efforts of risk management [154].

### Note 5

While our overview treats these as distinct from another for simplicity, note that hybrid [155] and multi-model approaches [156] have been developed. Due to our focus on mechanisms driving behavior change we do not address data-driven forecasting models in detail [see for example 157].

## Data Availability

No primary data were collected for this study. All data used in our statistical analyses are available to the public. References are provided throughout the manuscript and in the Supporting Information S1 and S2.

## Acknowledgements

The authors would like to thank Sophia Dietrich at UFZ for valuable support in the literature review for this analysis. We also thank Niels Wollschläger at UFZ for his advice and support in accessing and processing weather data.

## Supplementary Information 1: Statistical Analysis of Mobility Data

In the following, we present the models that support the results presented in Section 4.1. We specify our basis model as

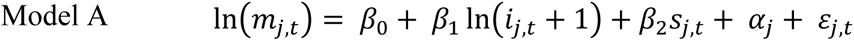

where *m*_*j*,*t*_ represents the dependent variable, the percentage change in mobility in federal state *j* on day *t*, relative to the average of the same month in the year 2019 [96]. Furthermore, *i*_*j*,*t*_represents the 7-day-incidence and *s*_*j*,*t*_ the stringency of containment measures in state *j* at day *t* [97, 98]. We calculate the natural log of incidence due to the at times exponential growth of case numbers and add one to address zeros in the data. α_*j*_ represents the individual fixed effect at the state level, ɛ_*j*,*t*_the error term. Note that we also dropped either predictor and tested whether including national incidence levels had a significant effect, as was the case in [13]. Neither resulted in an improved model fit.

As the raw data indicated significant changes in mobility patterns between weekdays, Saturdays and Sundays, we added individual indicator variables (*sat*_*t*_ & *sun*_*t*_) to account for this heterogeneity:

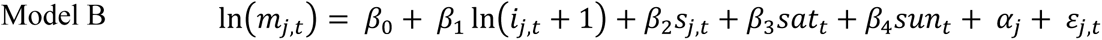

The weather changes in the fall likely impact mobility patterns. We include the daily average temperature *temp* and the daily average precipitation *precip* in state *j* at day *t*. This data was obtained from Deutscher Wetterdienst [158], Germany’s national meteorological service. The data for all 83 weather stations were downloaded and spatially interpolated for each federal state using the inverse distance weighting method.

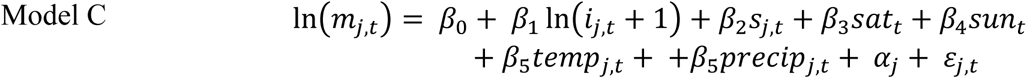

Finally, we estimate a model in which we drop the stringency index (*s*_*j*,*t*_) as a predictor variable and instead introduced a categorical variable *phase* which refers to the extent to which national-level NPIs were implemented and depends on the date of each observation. Any date before November 2 receives the value “local measures”, from November 2 to December 15 the value “lockdown light” and thereafter “lockdown hard”. This may also mitigate potential multicollinearity issues between *i*_*j*,*t*_and *s*_*j*,*t*_, which can occur if increased stringency follows increased incidence levels, as it was the case in some federal states. The model is thus specified as:

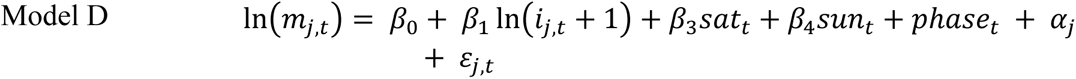

Note that daily the inclusion of weather data did not lead to improvements in model fit in the specification of Model D, perhaps because the different NPI phases roughly coincide with decreasing temperature levels and increased precipitation in fall. The variables *temp* and *precip* were thus not included in Model D.

Detailed regression results can be found in S1 Table 1. The results indicate that the sign of stringency and incidence are as expected and robust across all models, with slight reductions in effect sizes due to the incorporation of additional predictors. With declining temperature and higher precipitation mobility is reduced as should be expected. In Models A and B, this seasonal trend seemed to be attributed to increases in stringency and incidence over the same period. Interestingly, the introduction of the variable *phase* as an ordinal measure of stringency improved model fit while reducing the effect of incidence slightly, indicating that more variance in the data can be explained when measuring ordinal stringency at the national level. In Fig 4 in Section 4.1, a marginal effects plot for Model D is presented. Below, in S1 Fig 1, marginal effects for both state-level stringency and incidence in Model C are presented, depicting two values of the other predictor, and assuming a weekday mean values for temperature and precipitation.

**S1 Fig 1.**
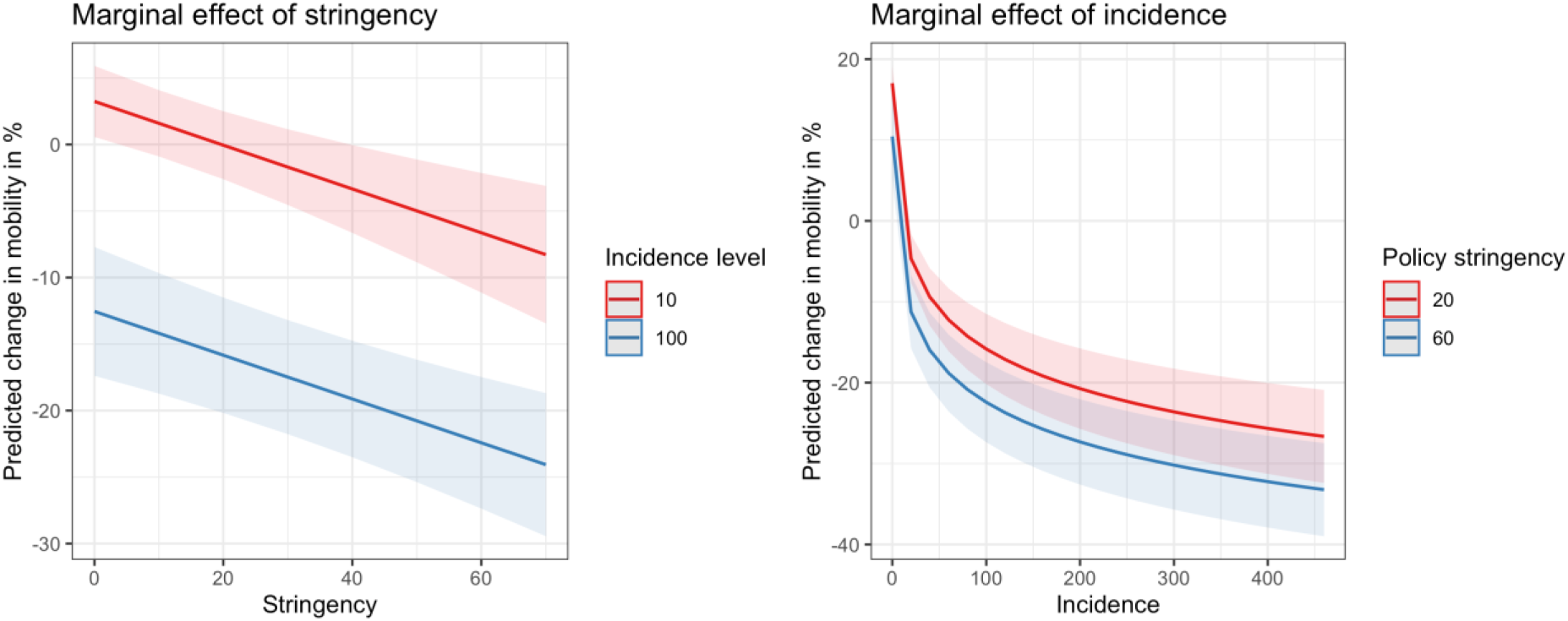
Marginal effects of policy stringency and 7-day incidence in Model C. The plot was generated using the R package *ggeffects* [99].

The models were subjected to diagnostic tests common for this model class: The Pesaran CD (Cross-Sectional Dependence) test indicated presence of heteroscedasticity and a Durbin-Watson test suggested presence of some serial correlation (see also the diagnostics plots in panels C and D of S1 Fig 2). We therefore report our regression results with standard errors robust to heteroscedasticity and autocorrelation, using the method of [159]. Models were estimated and standard errors calculated using the R package *fixest* [160]. As the models are implemented using a within transformation, multicollinearity that might have existed between time-related predictors and individual fixed effects is largely mitigated.

**S1 Fig 2.**
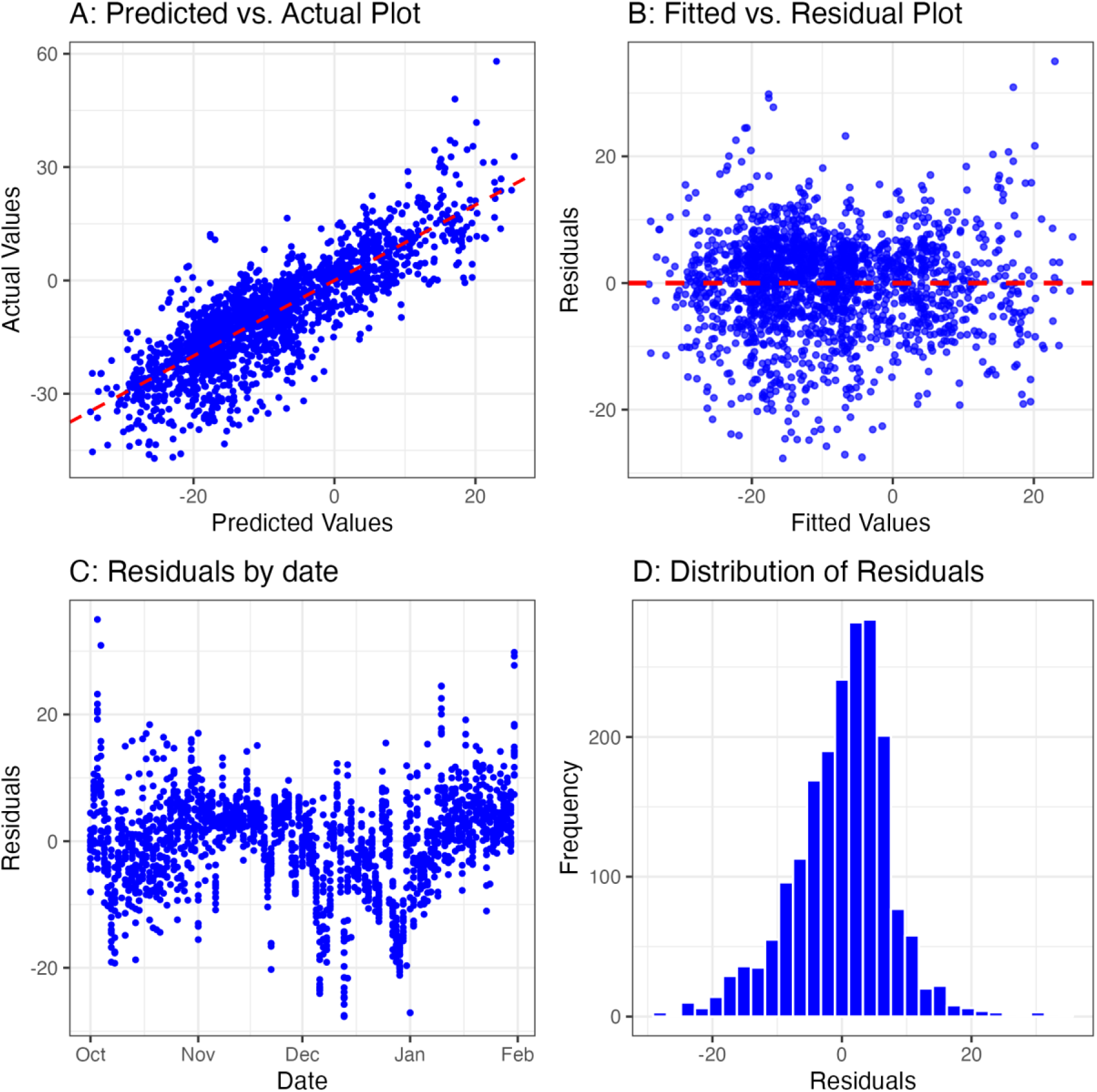
Model fit diagnostics for Model D.

**S1 Table 1.**
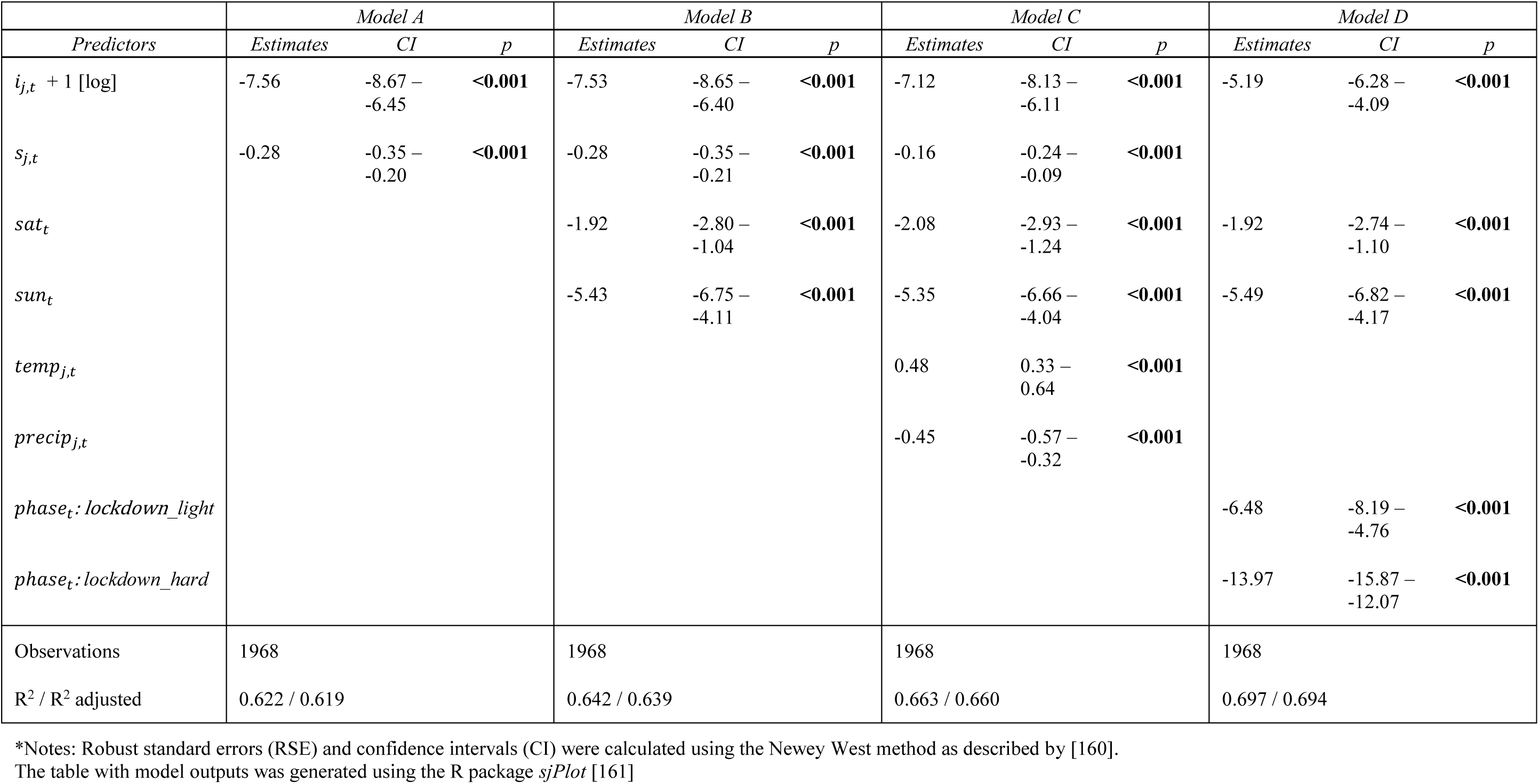
Results of fixed effects regression analyses.

## Supplementary Information 2: Statistical analysis of time variance

Here, we present the data and methodological approach supporting the results presented in Section 4.2.

### Data source

The data enabling the analysis in Section 4.2 stem from a representative survey of the German population dealing with perceptions and attitudes related to the COVID-19 pandemic [106]. The survey was implemented by the opinion research center *forsa* in 48 waves between March 18, 2020, and April, 27, 2022, by computer-assisted telephone interviews (size of the overall sample: n = 72,214). Respondents were randomly sampled from the German-speaking population aged 14 and above. Demographic information collected from survey participants includes sex, age, employment status, school-leaving qualification, household net income (grouped), preferences for the next federal election and past voting behavior. In the main survey, participants were asked to evaluate COVID-19 measures taken by the German government as well as other topics, varying with each wave. Frequently, this included questions on credibility of information provided on the pandemic by the German government and questions related to risk perception.

### Statistical analysis of diminishing risk perception

In three recurring questions of the survey, respondents were asked to rank the risk of infection for themselves, for their family members as well as the risk of spreading the disease to others. Surveyed individuals could respond to this question on a four-point rating scale. For our analysis of a potentially diminishing risk perception, these were matched with available data on 7-day incidence. The smallest possible geographic unit for matching both data was found to be the German state level. As no information on state of origin is available in [106] before August 2020, we only include data from thereafter. This results in 21 dates for which risk perception and incidence levels can be matched for each German state (n = 336). While the individual risk assessments in individual responses are on an ordinal scale, we assume that these can be treated as metric after calculating averages across the sample of each wave. To simplify the analysis and interpretation of results, we combine the data from three risk perception related questions into a composite variable. We calculate the arithmetic mean of the three variables for each state and date, which may also somewhat correct for the optimism bias common when merely the perceived infection risk for oneself is considered. Note, however, that we also repeated the analysis documented below for the three individual variables and the results proved robust, albeit with slight differences in effect sizes.

### Statistical models

Let *y*_*j*,*t*_represent the dependent variable, perceived risk, in state *j* at time *t*. Further, *x*_*j*,*t*_represents incidence level in state *j* at time *t* and, and *d*_*t*_ denotes a numeric representation of the date [162, 163]. For ease of interpretation, we convert this so that the first date in our data is represented by the number 1 and increases by 1 each day.

As a first step, we estimate a simple linear model (Model A):

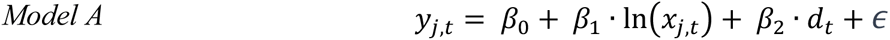

Note, however, that linear regression models have a number of relevant limitations with respect to time series due to, among other things, the assumption that observations are independent from one another [163]. We thus proceed to estimate two linear mixed effect models, which can handle clustered and hierarchical data in small sample sizes and provide more flexibility in modeling the data’s underlying structure [107]. In Model B and C, we assume incidence and the numeric time variable as fixed effects. Model B includes a state-level random effect *b*_*state*_ to account for unobserved heterogeneity among the 16 German federal states.

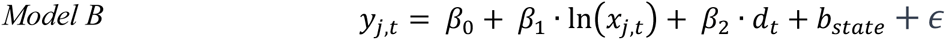

To investigate whether there are temporal patterns in the data beyond the linear progression of time (such as seasonal variations due to weather changes) we estimate a random effect *b*_*date*_, allowing the model intercept to vary for each observed date.

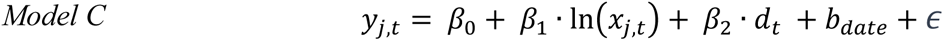

The results of the regression analyses are presented in S2 Table 1. Across all models, both predictor variables are significant and show the hypothesized sign: Perceived risk increases with incidence and decreases with the passage of time. While the effect size of *d*_*t*_may seem small at first glance, note that the distribution of the response variable is relatively small, with a mean of 2.14 (min: 1.50; max: 2.69), whereas *d*_*t*_ covers 610 days. In Fig 6 in Section 4.2.1, we present a partial effects plot depicting the impact of *β*_2_ over time at a given level of incidence. We used AIC to support model selection and find that the Model C indicates the overall best model fit, explaining about 60% of the variance in the data, with fixed effects accounting for 36%.

**S2 Table 1.**
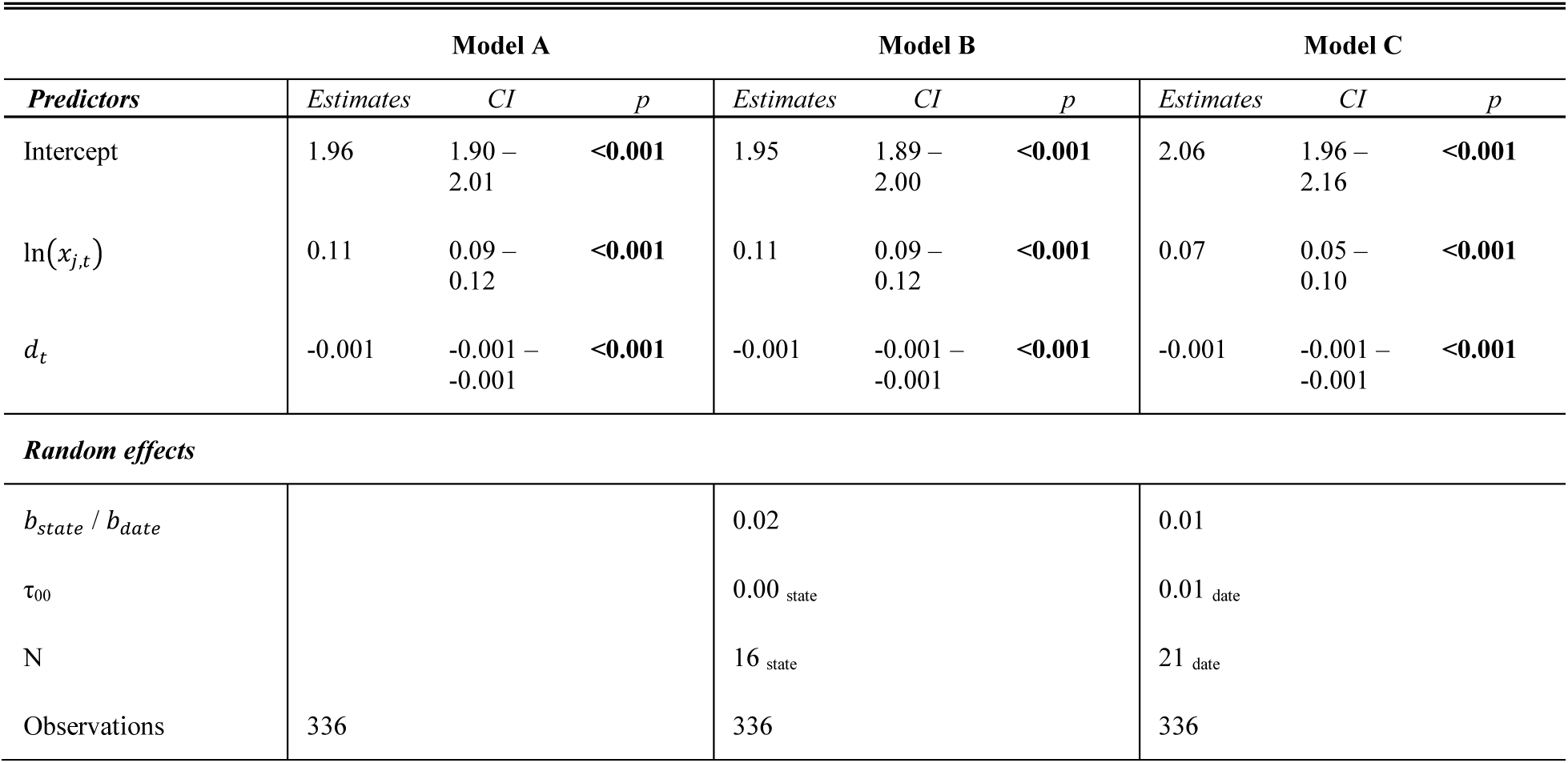
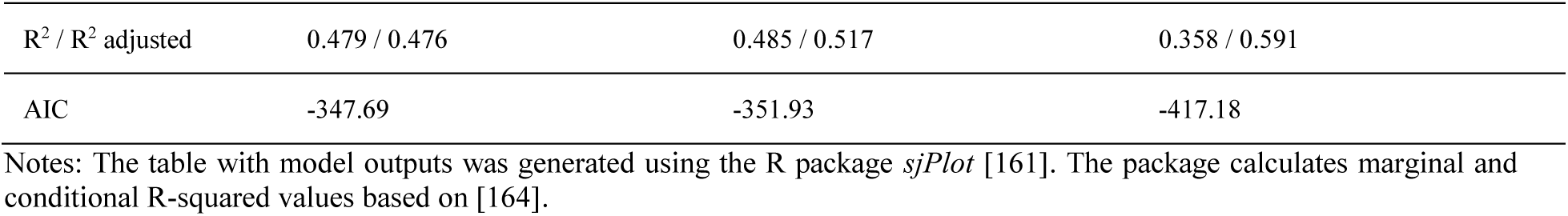
Regression results supporting **Section 4.2.1**.

#### Model diagnostic plots

Models B and C were subjected to standard goodness-of-fit tests for mixed effect models. We tested for multicollinearity through the calculation of variance inflation factors, which were found to be between 1.4 and 2.2 and thus in an acceptable range. In S2 Fig 1, we present a number of diagnostic plots for Model C that were used to assess the validity of all models.

**S2 Fig 1.**
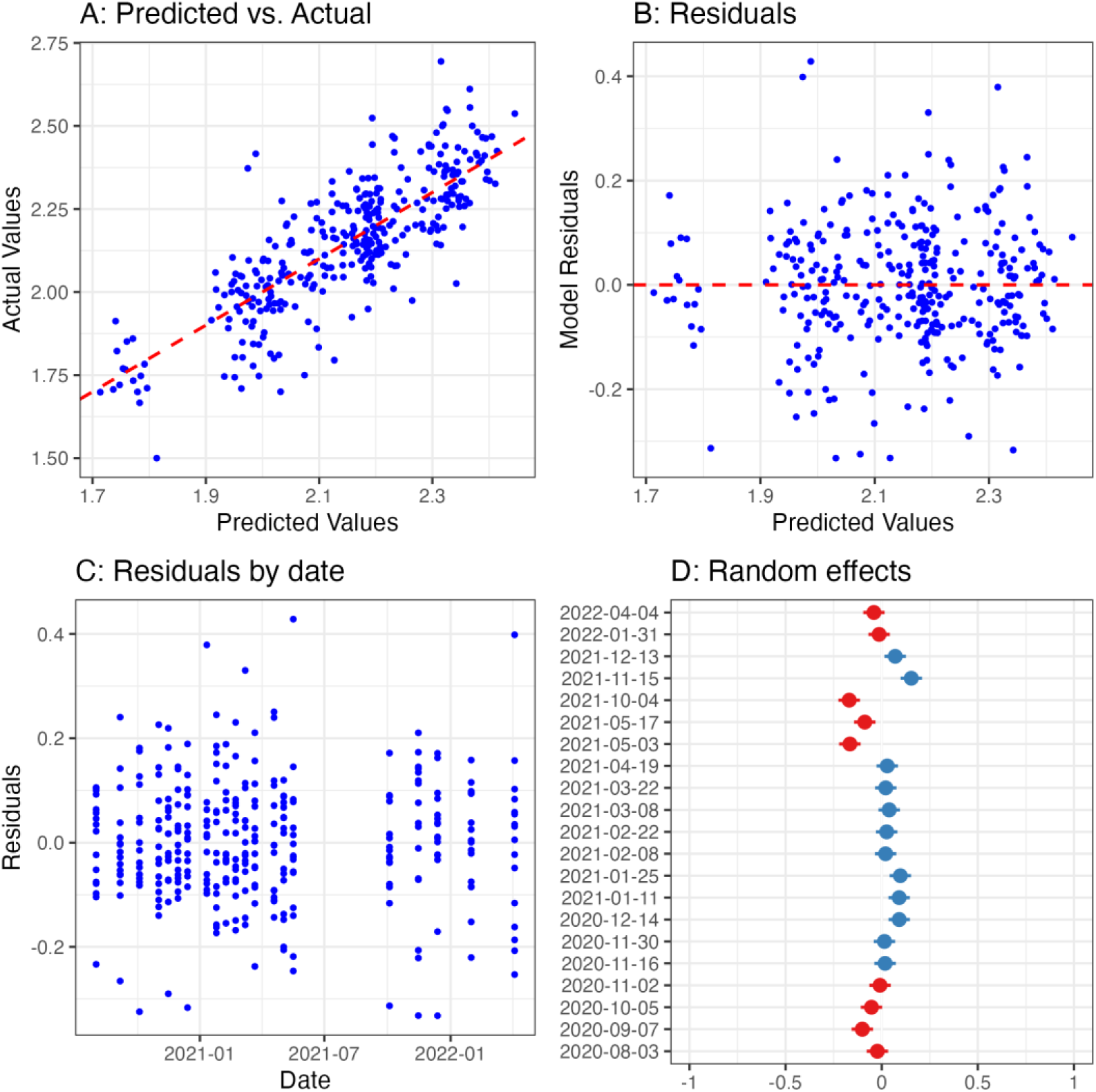
Diagnostic plots for Model C.

Panel A of S2 Fig 1 visualizes the overall model fit by plotting predicted against actual values. Panel B depicts model residuals, which do not show discernible patterns or signs of heteroscedasticity. In Panel C, the model residuals are plotted over the observed time period, exhibiting no visible pattern of temporal autocorrelation. Notably, the random effects plot in Panel D indicates a weak seasonal pattern introduced through the inclusion of date as a random effect, where the intercepts tend to increase slightly during most winter months.

### Statistical analysis of eroding trust and compliance

The data used in the analysis on trust in government information and assessment of containment measures also stem from [106]. All variables pertinent to the following analysis are listed in S2 Table 2, whereas the core interest is placed on (i) perceived *credibility* of government information and (ii) *assessment* of containment measures. Considering only survey waves for which data on both questions are available, 36 waves between April 2, 2020, and April 27, 2022, remain for analysis. For simplicity, we exclude responses from further analysis with the answer “I don’t know”, which correspond together with NA values to 2.0% and 0.9% for *assessment* and *credibility*, respectively.

As S2 Table 2 indicates, the data is collected through rating items with differing levels. For ordinal response variables, ordinal logistic regression methods are considered the standard and more robust than metric approaches [111, 165]. Traditional regression methods, however, incorporate ordinal *predictors* by treating these as nominal or numerical variables, with the risk of under- or overestimating their effects [166]. We thus employ a Bayesian approach using the R package *brms* [167], which allows for the inclusion of monotonic ordered predictors [166, 168]. We estimate two ordinal regression models with weakly informative priors, running four chains for 2,000 iterations. Algorithm convergence was confirmed through visual checks (“traceplots”) and the Rhat statistic. In the basic, univariate model, *assessment* is the dependent variable, with *credibility* as predictor. We extend this to a multivariate model by controlling for a number of socio-demographic variables. While we treat most other predictors as nominal, we also include income as a monotonic ordered predictor.

S2 Table 3 contains the summary of model results. It indicates that the thresholds for response variable categories as well as the effect of *credibility* are significant and robust across both model specification. The direction of effects is as expected: An increase in the credibility variable leads to an increase in assessment (note the levels of each variable in S2 Table 2). Another significant effect in the multivariate model is age above 60 years, which is consistent with this age group having a higher risk of mortality and thus less inclination to consider containment measures “go too far”. A comparison of both models was carried out using the leave-one-out information criterion (LOOIC), a Bayesian information criterion based on out-of-sample predictive performance. The comparison of LOOIC values (see S2 Table 3) indicated that the multivariate model has an overall better model fit, with a difference of more than two standard errors, indicating substantial improvement in large data sets [111]. We thus use this model to develop the conditional effect plot (Fig 8) presented in Section 4.2.2.

**S2 Table 2.**
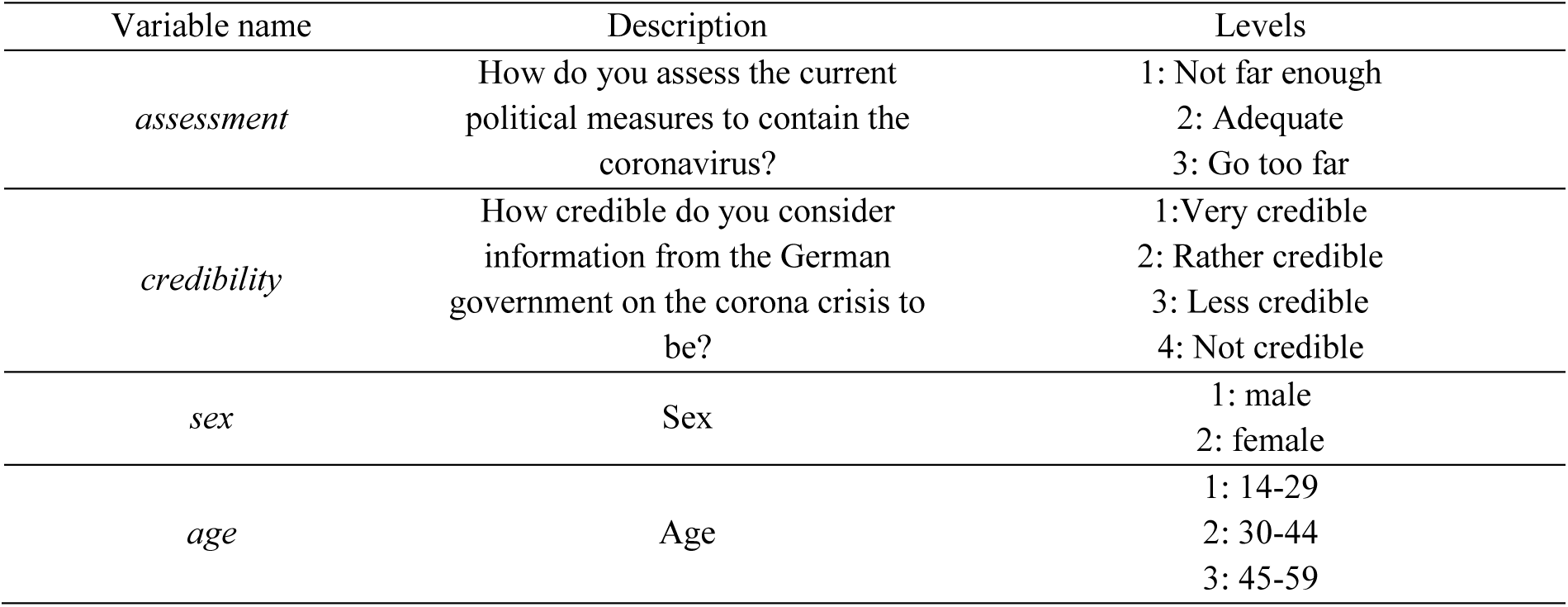
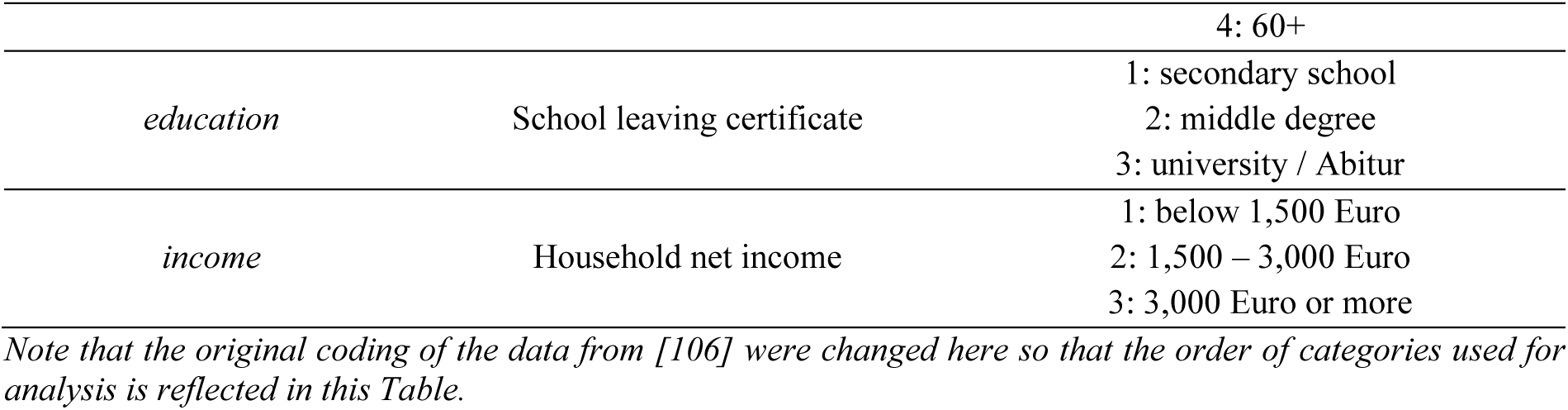
Model variables: Government credibility and assessment of response.

**S2 Table 3.**
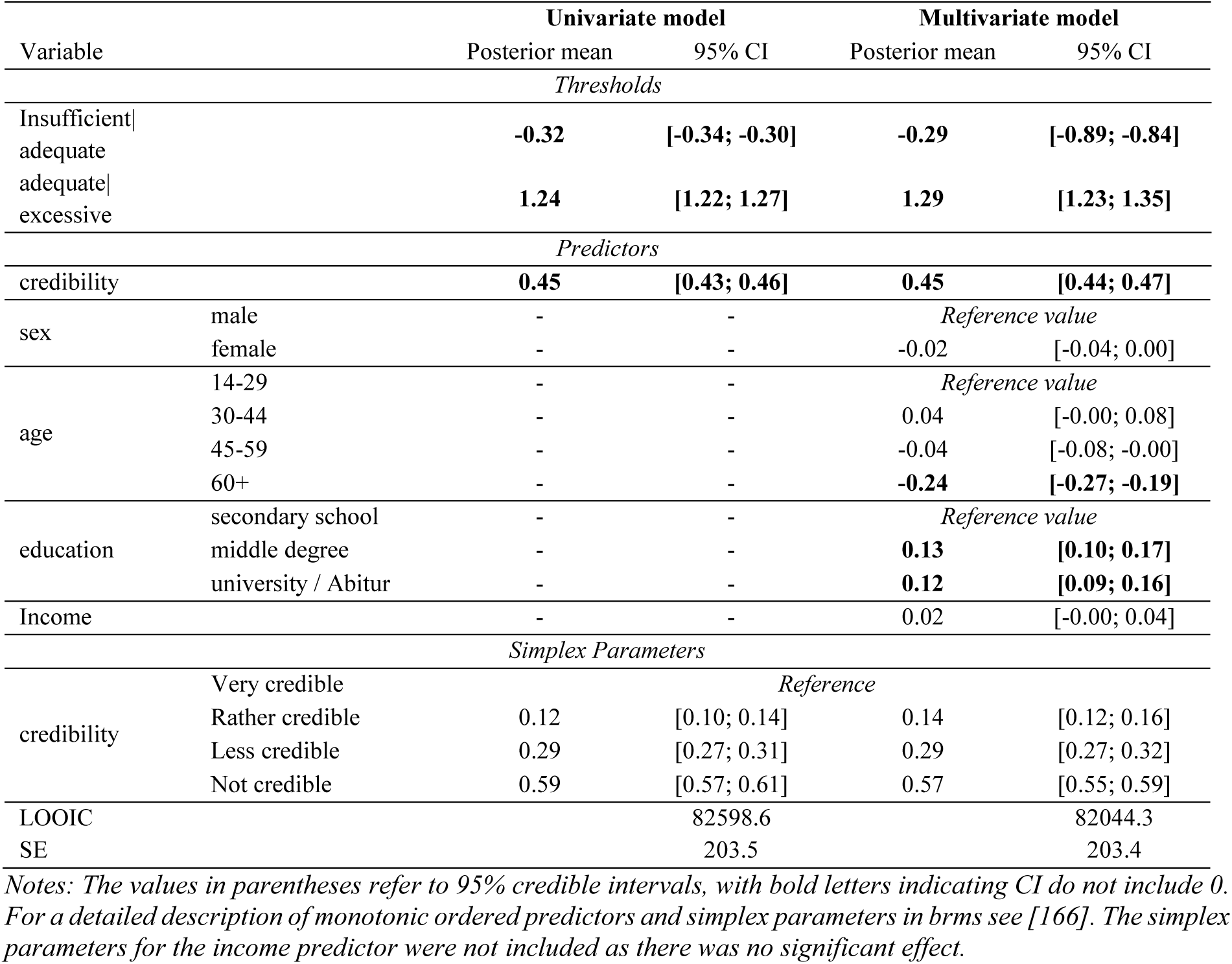
Results of ordinal regression.

## Supplementary Information 3: Conceptual SIR model of autonomous and policy-induced adaptation

The conceptual model used to support the discussion in Section 5 and generate Figs 10 and 11 builds on the classic Susceptible-Infected-Recovered (SIR) model [146]. The model is formulated as:

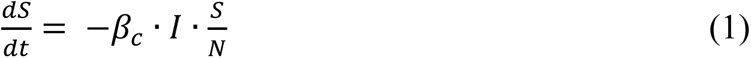

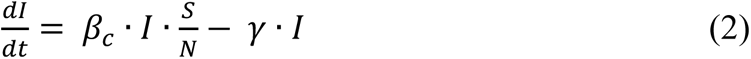

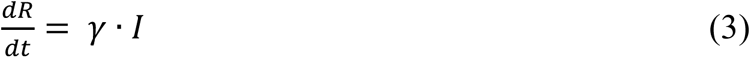

where S denotes the susceptible population, I the infected population, R the stock of removed population (either by death or recovery), and N the total population. Deviating from the classic SIR model, we assume a time-varying transmission rate *β*_*c*_, defined as:

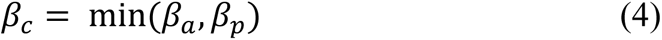

As explained in Section 5, we assume an overlapping effect of autonomous and policy-induced adaptation in *β*_*c*_, which is implemented as the minimum of the hypothetical transmission rates *β*_*a*_, denoting endogenous behavioral response without considering impacts of NPIs, and *β*_*p*_, denoting policy-induced changes in contacts without considering endogenous behavioral response.

### Autonomous adaptation

To define the impact of autonomous adaptation *β*_*a*_, we follow an existing application [24] and assume that individuals derive utility *u*(*β*_*a*_) from social contacts, specified as:

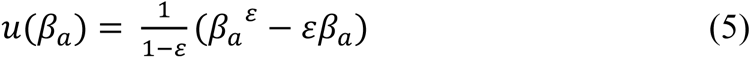

In (5), the parameter ε ∈ (0,1) captures how important it is for individuals to engage in physical contacts. Assuming an early pandemic situation, where still almost all of the population is susceptible, the individual risk of an infection is β I S/N ≈ β I. As described in detail in [24], a rational, risk-averse individual chooses the contacts such as to trade-off current utility from contacts, (5), and the expected utility loss from an infection, Δv. Following from this, the number of contacts is determined by

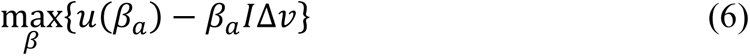

With the specified utility function (5), the optimal number of contacts becomes a decreasing function of the number of infected, I.

The first-order condition for (6) reads

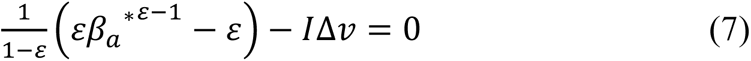

which can be rearranged to

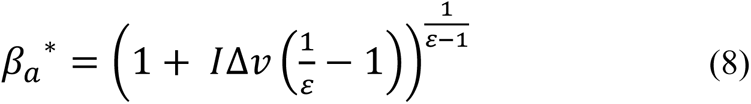

### Policy-induced adaptation

The effect of NPIs on contacts and transmissions is introduced in the model as a direct reduction of contacts. The transmission rate *β*_*p*_ is set by a piecewise constant function:

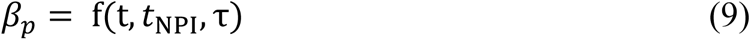

where t denotes the current time step, *t*_NPI_ denotes the time step when NPIs are introduced and the parameter τ denotes the value to which *β*_*p*_ is set in a smoothed jump over seven days for starting from t = *t*_NPI_.

### Parameters

Due to the illustrative function of the model, parameter values were set deliberately, as specified in S3 Table 1.

**S3 Table 1.**
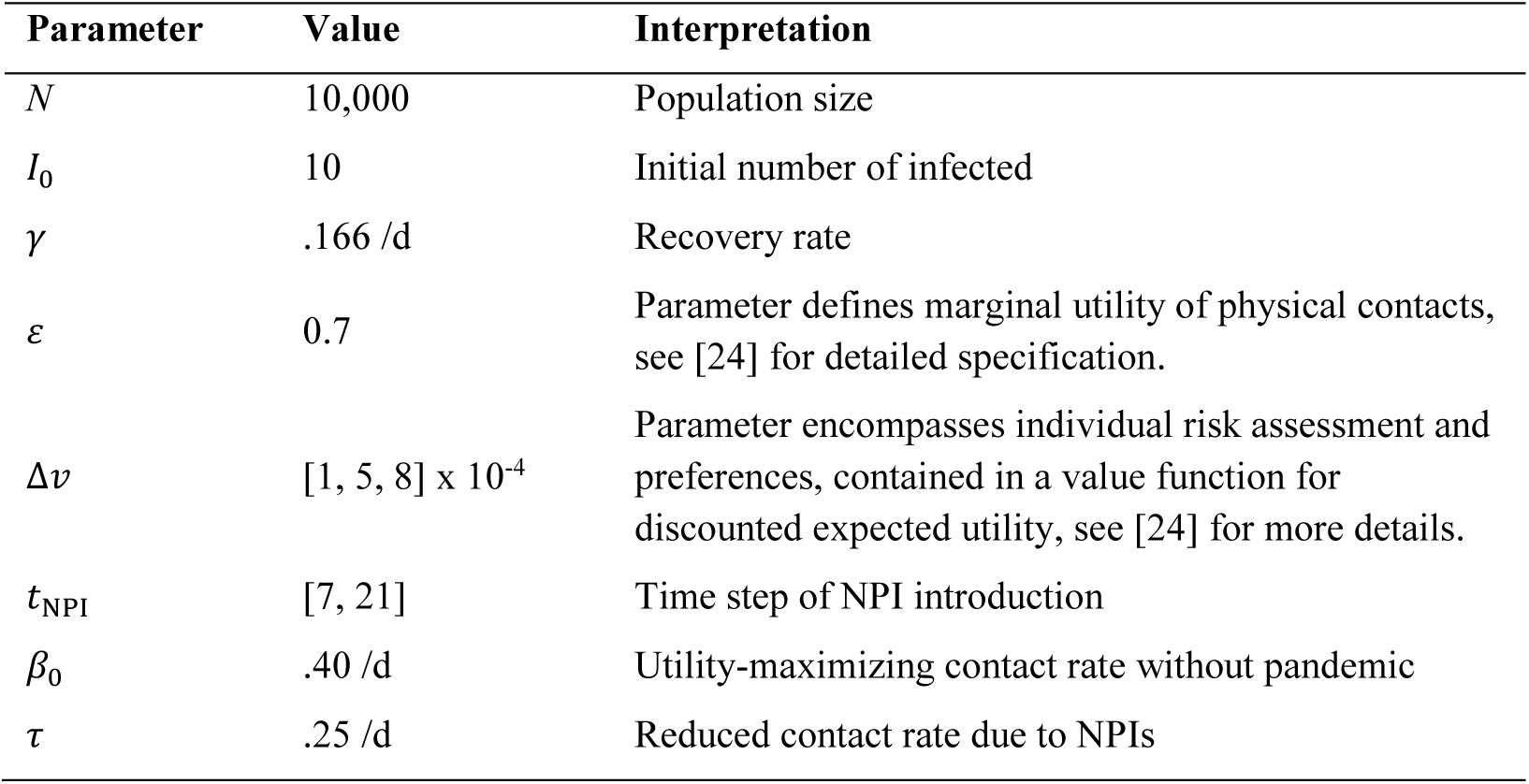
Model parameters.

