## Supplementary Information 1 for "Autonomous and policy-induced behavior change during the COVID-19 pandemic: Towards understanding and modeling the interplay of behavioral adaptation"

### Supplementary Information 1: Statistical Analysis of Mobility Data

In the following, we present the models that support the results presented in Section 4.1. We specify our basis model as

Model A $$\ln(m_{j,t}) = \beta_0 + \beta_1 \ln(i_{j,t} + 1) + \beta_2 s_{j,t} + \alpha_j + \varepsilon_{j,t}$$

where $m_{j,t}$ represents the dependent variable, the percentage change in mobility in federal state *j* on day *t*, relative to the average of the same month in the year 2019 [1]. Furthermore, $i_{j,t}$ represents the 7-day-incidence and $s_{j,t}$ the stringency of containment measures in state *j* at day *t* [2, 3]. We calculate the natural log of incidence due to the at times exponential growth of case numbers and add one to address zeros in the data. $\alpha_j$ represents the individual fixed effect at the state level, $\varepsilon_{j,t}$ the error term. Note that we also dropped either predictor and tested whether including national incidence levels had a significant effect, as was the case in [4]. Neither resulted in an improved model fit.

As the raw data indicated significant changes in mobility patterns between weekdays, Saturdays and Sundays, we added individual indicator variables ($sat_t$ & $sun_t$) to account for this heterogeneity:

Model B $$\ln(m_{j,t}) = \beta_0 + \beta_1 \ln(i_{j,t} + 1) + \beta_2 s_{j,t} + \beta_3 sat_t + \beta_4 sun_t + \alpha_j + \varepsilon_{j,t}$$

The weather changes in the fall likely impact mobility patterns. We include the daily average temperature *temp* and the daily average precipitation *precip* in state *j* at day *t*. This data was obtained from Deutscher Wetterdienst [5], Germany's national meteorological service. The data for all 83 weather stations were downloaded and spatially interpolated for each federal state using the inverse distance weighting method.

Model C $$\ln(m_{j,t}) = \beta_0 + \beta_1 \ln(i_{j,t} + 1) + \beta_2 s_{j,t} + \beta_3 sat_t + \beta_4 sun_t + \beta_5 temp_{j,t} + +\beta_5 precip_{j,t} + \alpha_j + \varepsilon_{j,t}$$

Finally, we estimate a model in which we drop the stringency index ($s_{j,t}$) as a predictor variable and instead introduced a categorical variable *phase* which refers to the extent to which national-level NPIs were implemented and depends on the date of each observation. Any date before November 2 receives the value "local measures", from November 2 to December 15 the value "lockdown light" and thereafter "lockdown hard". This may also mitigate potential multicollinearity issues between $i_{j,t}$ and $s_{j,t}$, which can occur if increased stringency follows increased incidence levels, as it was the case in some federal states. The model is thus specified as:

Model D $$\ln(m_{j,t}) = \beta_0 + \beta_1 \ln(i_{j,t} + 1) + \beta_3 sat_t + \beta_4 sun_t + phase_t + \alpha_j + \varepsilon_{j,t}$$

Note that daily the inclusion of weather data did not lead to improvements in model fit in the specification of Model D, perhaps because the different NPI phases roughly coincide with decreasing temperature levels and increased precipitation in fall. The variables *temp* and *precip* were thus not included in Model D.

Detailed regression results can be found in S1 Table 1. The results indicate that the sign of stringency and incidence are as expected and robust across all models, with slight reductions in effect sizes due to the incorporation of additional predictors. With declining temperature and higher precipitation mobility is reduced as should be expected. In Models A and B, this seasonal trend seemed to be attributed to increases in stringency and incidence over the same period. Interestingly, the introduction of the variable *phase* as an ordinal measure of stringency improved model fit while reducing the effect of incidence slightly, indicating that more variance in the data can be explained when measuring ordinal stringency at the national level. In Fig 4 in Section 4.1, a marginal effects plot for Model D is presented. Below, in S1 Fig 1, marginal effects for both state-level stringency and incidence in Model C are presented, depicting two values of the other predictor, and assuming a weekday mean values for temperature and precipitation.

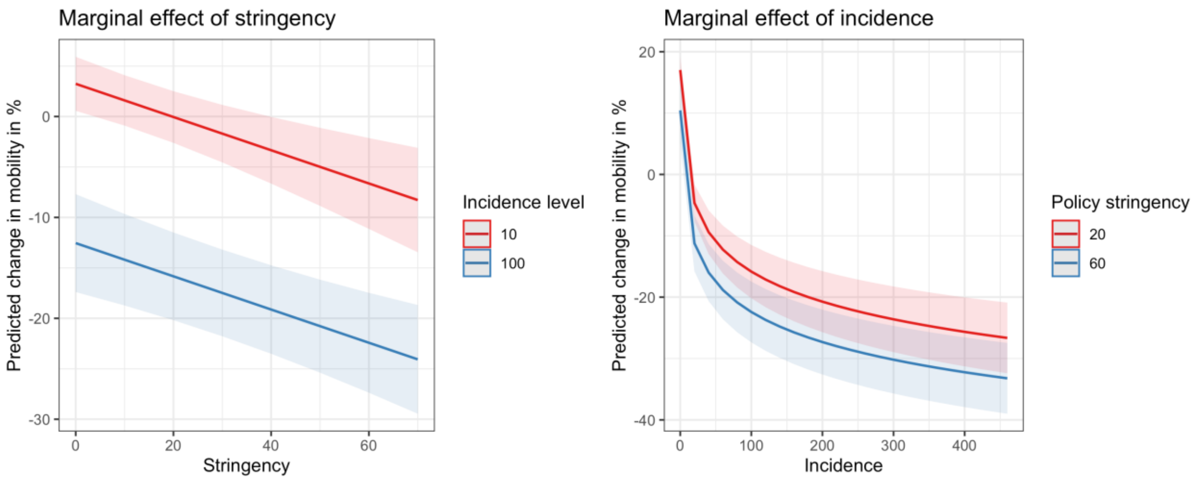


**S1 Fig 1. Marginal effects of policy stringency and 7-day incidence in Model C.** The plot was generated using the R package *ggeffects* [6].

The models were subjected to diagnostic tests common for this model class: The Pesaran CD (Cross-Sectional Dependence) test indicated presence of heteroscedasticity and a Durbin-Watson test suggested presence of some serial correlation (see also the diagnostics plots in panels C and D of S1 Fig 2). We therefore report our regression results with standard errors robust to heteroscedasticity and autocorrelation, using the method of [7]. Models were estimated and standard errors calculated using the R package *fixest* [8]. As the models are implemented using a within transformation, multicollinearity that might have existed between time-related predictors and individual fixed effects is largely mitigated.

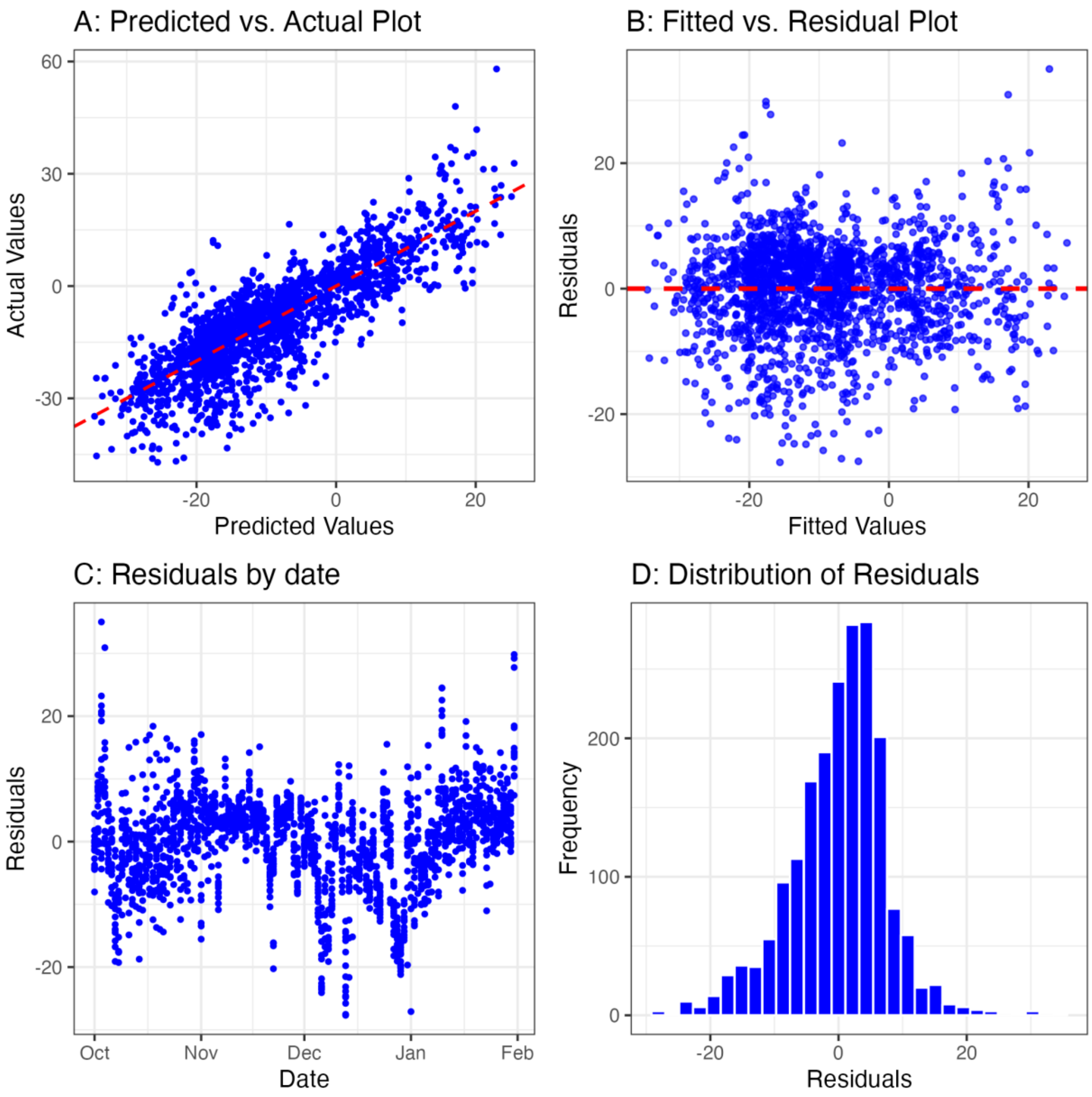


**S1 Fig 2. Model fit diagnostics for Model D.**

#### S1 Table 1. Results of fixed effects regression analyses.

| | *Model A* | | | *Model B* | | | *Model C* | | | *Model D* | | |
|---|---|---|---|---|---|---|---|---|---|---|---|---|
| *Predictors* | *Estimates* | *CI* | *p* | *Estimates* | *CI* | *p* | *Estimates* | *CI* | *p* | *Estimates* | *CI* | *p* |
| $i_{j,t}$ + 1 [log] | -7.56 | -8.67 – -6.45 | **<0.001** | -7.53 | -8.65 – -6.40 | **<0.001** | -7.12 | -8.13 – -6.11 | **<0.001** | -5.19 | -6.28 – -4.09 | **<0.001** |
| $s_{j,t}$ | -0.28 | -0.35 – -0.20 | **<0.001** | -0.28 | -0.35 – -0.21 | **<0.001** | -0.16 | -0.24 – -0.09 | **<0.001** | | | |
| $sat_t$ | | | | -1.92 | -2.80 – -1.04 | **<0.001** | -2.08 | -2.93 – -1.24 | **<0.001** | -1.92 | -2.74 – -1.10 | **<0.001** |
| $sun_t$ | | | | -5.43 | -6.75 – -4.11 | **<0.001** | -5.35 | -6.66 – -4.04 | **<0.001** | -5.49 | -6.82 – -4.17 | **<0.001** |
| $temp_{j,t}$ | | | | | | | 0.48 | 0.33 – 0.64 | **<0.001** | | | |
| $precip_{j,t}$ | | | | | | | -0.45 | -0.57 – -0.32 | **<0.001** | | | |
| $phase_t$*: lockdown_light* | | | | | | | | | | -6.48 | -8.19 – -4.76 | **<0.001** |
| $phase_t$*: lockdown_hard* | | | | | | | | | | -13.97 | -15.87 – -12.07 | **<0.001** |
| Observations | 1968 | | | 1968 | | | 1968 | | | 1968 | | |
| $R^2$ / $R^2$ adjusted | 0.622 / 0.619 | | | 0.642 / 0.639 | | | 0.663 / 0.660 | | | 0.697 / 0.694 | | |

*Notes: Robust standard errors (RSE) and confidence intervals (CI) were calculated using the Newey West method as described by [8].
The table with model outputs was generated using the R package *sjPlot* [9]
