## Supplementary Information 2 for "Autonomous and policy-induced behavior change during the COVID-19 pandemic: Towards understanding and modeling the interplay of behavioral adaptation"

As a first step, we estimate a simple linear model (Model A):

*Model A* $$y_{j,t} = \beta_0 + \beta_1 \cdot \ln(x_{j,t}) + \beta_2 \cdot d_t + \epsilon$$

Note, however, that linear regression models have a number of relevant limitations with respect to time series due to, among other things, the assumption that observations are independent from one another [3]. We thus proceed to estimate two linear mixed effect models, which can handle clustered and hierarchical data in small sample sizes and provide more flexibility in modeling the data's underlying structure [4]. In Model B and C, we assume incidence and the numeric time variable as fixed effects. Model B includes a state-level random effect $b_{state}$ to account for unobserved heterogeneity among the 16 German federal states.

*Model B* $$y_{j,t} = \beta_0 + \beta_1 \cdot \ln(x_{j,t}) + \beta_2 \cdot d_t + b_{state} + \epsilon$$

To investigate whether there are temporal patterns in the data beyond the linear progression of time (such as seasonal variations due to weather changes) we estimate a random effect $b_{date}$, allowing the model intercept to vary for each observed date.

*Model C* $$y_{j,t} = \beta_0 + \beta_1 \cdot \ln(x_{j,t}) + \beta_2 \cdot d_t + b_{date} + \epsilon$$

The results of the regression analyses are presented in S2 Table 1. Across all models, both predictor variables are significant and show the hypothesized sign: Perceived risk increases with incidence and decreases with the passage of time. While the effect size of $d_t$ may seem small at first glance, note that the distribution of the response variable is relatively small, with a mean of 2.14 (min: 1.50; max: 2.69), whereas $d_t$ covers 610 days. In Fig 6 in Section 4.2.1, we present a partial effects plot depicting the impact of $\beta_2$ over time at a given level of incidence. We used AIC to support model selection and find that the Model C indicates the overall best model fit, explaining about 60% of the variance in the data, with fixed effects accounting for 36%.

**S2 Table 1. Regression results supporting Section 4.2.1**

| | **Model A** | | | **Model B** | | | **Model C** | | |
|---|---|---|---|---|---|---|---|---|---|
| ***Predictors*** | *Estimates* | *CI* | *p* | *Estimates* | *CI* | *p* | *Estimates* | *CI* | *p* |
| Intercept | 1.96 | 1.90 – 2.01 | **<0.001** | 1.95 | 1.89 – 2.00 | **<0.001** | 2.06 | 1.96 – 2.16 | **<0.001** |
| $\ln(x_{j,t})$ | 0.11 | 0.09 – 0.12 | **<0.001** | 0.11 | 0.09 – 0.12 | **<0.001** | 0.07 | 0.05 – 0.10 | **<0.001** |
| $d_t$ | -0.001 | -0.001 – -0.001 | **<0.001** | -0.001 | -0.001 – -0.001 | **<0.001** | -0.001 | -0.001 – -0.001 | **<0.001** |
| ***Random effects*** | | | | | | | | | |
| $b_{state}$ / $b_{date}$ | | | | 0.02 | | | 0.01 | | |
| $\tau_{00}$ | | | | 0.00 $_{state}$ | | | 0.01 $_{date}$ | | |
| N | | | | 16 $_{state}$ | | | 21 $_{date}$ | | |
| Observations | 336 | | | 336 | | | 336 | | |

| | | | |
|---|---|---|---|
| $R^2$ / $R^2$ adjusted | 0.479 / 0.476 | 0.485 / 0.517 | 0.358 / 0.591 |
| AIC | -347.69 | -351.93 | -417.18 |

Notes: The table with model outputs was generated using the R package *sjPlot* [5]. The package calculates marginal and conditional R-squared values based on [6].

### *Model diagnostic plots*

Models B and C were subjected to standard goodness-of-fit tests for mixed effect models. We tested for multicollinearity through the calculation of variance inflation factors, which were found to be between 1.4 and 2.2 and thus in an acceptable range. In S2 Fig 1, we present a number of diagnostic plots for Model C that were used to assess the validity of all models.

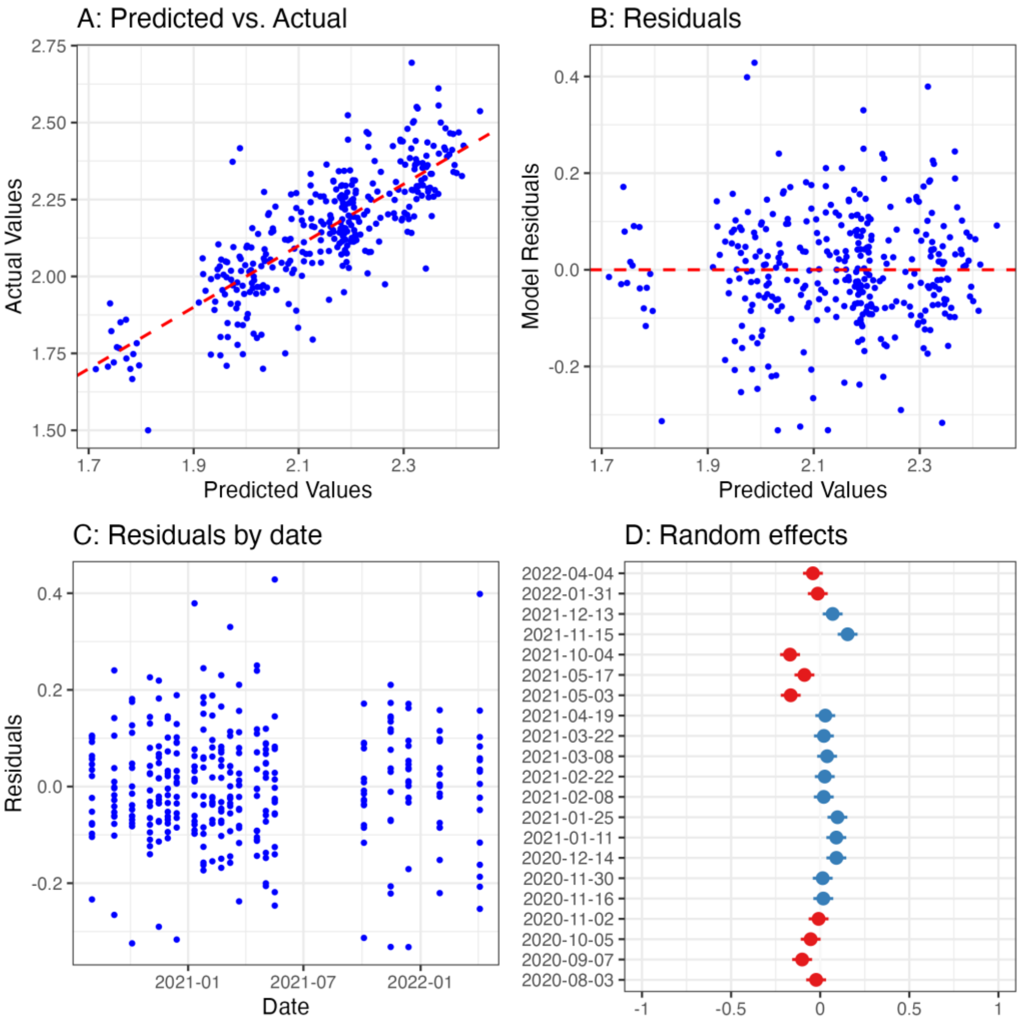


**S2 Fig 1. Diagnostic plots for Model C**

Panel A of S2 Fig 1 visualizes the overall model fit by plotting predicted against actual values. Panel B depicts model residuals, which do not show discernible patterns or signs of heteroscedasticity. In Panel C, the model residuals are plotted over the observed time period, exhibiting no visible pattern of temporal autocorrelation. Notably, the random effects plot in Panel D indicates a weak seasonal pattern introduced through the inclusion of date as a random effect, where the intercepts tend to increase slightly during most winter months.

**S2 Table 2. Model variables: Government credibility and assessment of response**.

| Variable name | Description | Levels |
|---|---|---|
| *assessment* | How do you assess the current political measures to contain the coronavirus? | 1: Not far enough<br>2: Adequate<br>3: Go too far |
| *credibility* | How credible do you consider information from the German government on the corona crisis to be? | 1:Very credible<br>2: Rather credible<br>3: Less credible<br>4: Not credible |
| *sex* | Sex | 1: male<br>2: female |
| *age* | Age | 1: 14-29<br>2: 30-44<br>3: 45-59 |

| | | |
|---|---|---|
| | | 4: 60+ |
| *education* | School leaving certificate | 1: secondary school<br>2: middle degree<br>3: university / Abitur |
| *income* | Household net income | 1: below 1,500 Euro<br>2: 1,500 – 3,000 Euro<br>3: 3,000 Euro or more |

*Note that the original coding of the data from [1] were changed here so that the order of categories used for analysis is reflected in this Table.*

### S2 Table 3. Results of ordinal regression.

| | | **Univariate model** | | **Multivariate model** | |
|---|---|---|---|---|---|
| Variable | | Posterior mean | 95% CI | Posterior mean | 95% CI |
| *Thresholds* | | | | | |
| Insufficient\| adequate | | **-0.32** | **[-0.34; -0.30]** | **-0.29** | **[-0.89; -0.84]** |
| adequate\| excessive | | **1.24** | **[1.22; 1.27]** | **1.29** | **[1.23; 1.35]** |
| *Predictors* | | | | | |
| credibility | | **0.45** | **[0.43; 0.46]** | **0.45** | **[0.44; 0.47]** |
| sex | male | - | - | *Reference value* | |
| | female | - | - | -0.02 | [-0.04; 0.00] |
| age | 14-29 | - | - | *Reference value* | |
| | 30-44 | - | - | 0.04 | [-0.00; 0.08] |
| | 45-59 | - | - | -0.04 | [-0.08; -0.00] |
| | 60+ | - | - | **-0.24** | **[-0.27; -0.19]** |
| education | secondary school | - | - | *Reference value* | |
| | middle degree | - | - | **0.13** | **[0.10; 0.17]** |
| | university / Abitur | - | - | **0.12** | **[0.09; 0.16]** |
| Income | | - | - | 0.02 | [-0.00; 0.04] |
| *Simplex Parameters* | | | | | |
| credibility | Very credible | | *Reference* | | |
| | Rather credible | 0.12 | [0.10; 0.14] | 0.14 | [0.12; 0.16] |
| | Less credible | 0.29 | [0.27; 0.31] | 0.29 | [0.27; 0.32] |
| | Not credible | 0.59 | [0.57; 0.61] | 0.57 | [0.55; 0.59] |
| LOOIC | | | 82598.6 | | 82044.3 |
| SE | | | 203.5 | | 203.4 |

*Notes: The values in parentheses refer to 95% credible intervals, with bold letters indicating CI do not include 0. For a detailed description of monotonic ordered predictors and simplex parameters in brms see [9]. The simplex parameters for the income predictor were not included as there was no significant effect.*
