## Supplementary Information 3 for "Autonomous and policy-induced behavior change during the COVID-19 pandemic: Towards understanding and modeling the interplay of behavioral adaptation"

### Supplementary Information 3: Conceptual SIR model of autonomous and policy-induced adaptation

The conceptual model used to support the discussion in Section 5 and generate Figs 10 and 11 builds on the classic Susceptible-Infected-Recovered (SIR) model [1]. The model is formulated as:

$$\frac{dS}{dt} = -\beta_c \cdot I \cdot \frac{S}{N} \tag{1}$$

$$\frac{dI}{dt} = \beta_c \cdot I \cdot \frac{S}{N} - \gamma \cdot I \tag{2}$$

$$\frac{dR}{dt} = \gamma \cdot I \tag{3}$$

where S denotes the susceptible population, I the infected population, R the stock of removed population (either by death or recovery), and N the total population. Deviating from the classic SIR model, we assume a time-varying transmission rate $\beta_c$, defined as:

$$\beta_c = \min(\beta_a, \beta_p) \tag{4}$$

As explained in Section 5, we assume an overlapping effect of autonomous and policy-induced adaptation in $\beta_c$, which is implemented as the minimum of the hypothetical transmission rates $\beta_a$, denoting endogenous behavioral response without considering impacts of NPIs, and $\beta_p$, denoting policy-induced changes in contacts without considering endogenous behavioral response.

#### Autonomous adaptation

To define the impact of autonomous adaptation $\beta_a$, we follow an existing application [2] and assume that individuals derive utility $u(\beta_a)$ from social contacts, specified as:

$$u(\beta_a) = \frac{1}{1-\varepsilon}({\beta_a}^{\varepsilon} - \varepsilon\beta_a) \tag{5}$$

In (5), the parameter $\varepsilon \in (0,1)$ captures how important it is for individuals to engage in physical contacts. Assuming an early pandemic situation, where still almost all of the population is susceptible, the individual risk of an infection is $\beta$ I S/N $\approx$ $\beta$ I. As described in detail in [2], a rational, risk-averse individual chooses the contacts such as to trade-off current utility from contacts, (5), and the expected utility loss from an infection, $\Delta$v. Following from this, the number of contacts is determined by

$$\max_{\beta}\{u(\beta_a) - \beta_a I \Delta v\} \tag{6}$$

With the specified utility function (5), the optimal number of contacts becomes a decreasing function of the number of infected, I.

The first-order condition for (6) reads

$$\frac{1}{1-\varepsilon}\left(\varepsilon {\beta_a}^{*\varepsilon-1} - \varepsilon\right) - I\Delta v = 0 \tag{7}$$

which can be rearranged to

$${\beta_a}^{*} = \left(1 + I\Delta v\left(\frac{1}{\varepsilon} - 1\right)\right)^{\frac{1}{\varepsilon-1}} \tag{8}$$

#### Policy-induced adaptation

The effect of NPIs on contacts and transmissions is introduced in the model as a direct reduction of contacts. The transmission rate $\beta_p$ is set by a piecewise constant function:

$$\beta_p = \mathrm{f}(\mathrm{t}, t_{\mathrm{NPI}}, \tau) \tag{9}$$

where t denotes the current time step, $t_{\mathrm{NPI}}$ denotes the time step when NPIs are introduced and the parameter τ denotes the value to which $\beta_p$ is set in a smoothed jump over seven days for starting from t = $t_{\mathrm{NPI}}$.

#### Parameters

Due to the illustrative function of the model, parameter values were set deliberately, as specified in S3 Table 1.

**S3 Table 1. Model parameters.**

| Parameter | Value | Interpretation |
|---|---|---|
| $N$ | 10,000 | Population size |
| $I_0$ | 10 | Initial number of infected |
| $\gamma$ | .166 /d | Recovery rate |
| $\varepsilon$ | 0.7 | Parameter defines marginal utility of physical contacts, see [2] for detailed specification. |
| $\Delta v$ | [1, 5, 8] x $10^{-4}$ | Parameter encompasses individual risk assessment and preferences, contained in a value function for discounted expected utility, see [2] for more details. |
| $t_{\mathrm{NPI}}$ | [7, 21] | Time step of NPI introduction |
| $\beta_0$ | .40 /d | Utility-maximizing contact rate without pandemic |
| $\tau$ | .25 /d | Reduced contact rate due to NPIs |
